# Performance characteristics and potential public health impact of improved pre-erythrocytic malaria vaccines targeting childhood burden

**DOI:** 10.1101/2024.11.12.24317082

**Authors:** Josephine Malinga, Lydia Braunack-Mayer, Thiery Masserey, Aurélien Cavelan, Daniel Chandramohan, Alassane Dicko, Jean-Bosco Ouédraogo, Sherrie L Kelly, Epke A Le Rutte, Narimane Nekkab, Melissa A Penny

## Abstract

New malaria vaccine development builds on groundbreaking recommendations and roll-out of two approved pre-erythrocytic vaccines (PEVs); RTS,S/AS01 and R21/MM. Whilst these vaccines are effective in reducing childhood malaria within yearly routine immunization programs or seasonal vaccination, there is little evidence on how different PEV efficacies, durations of protection, and spacing between doses influence the potential to avert uncomplicated and severe childhood malaria. Mainly, lacking understanding of the required vaccine properties and delivery strategies that lead to an effective childhood vaccine with multi-year protection. We used an individual-based model of malaria transmission informed by trial data to quantify trade-offs between PEV performance properties and impact across different endemicities, deployment schedules, and coverage levels.

We found that deploying a vaccine with 90% initial efficacy, with a six to 12-month half-life duration of protection, co-administered with a blood-stage drug, followed by yearly boosters, results in 60-80% yearly incidence reduction, consistent with seasonal RTS,S and R21 trials. Halting vaccination after five years, leads to sustained protection of at least a 35% incidence reduction in children <six years in the 12 months following cessation in settings where *Pf*PR_2-10_ <30%. Increasing the half-life duration to 12 -18 months or reaching more children provides the same health impact with lower vaccine efficacy. Without a booster (fourth dose), high efficacy (>90%) and longer half-life duration (>12 months) are required to sustain impact beyond primary vaccination, averting up to half the preceding year’s burden. The contribution of each property to the overall impact varies by setting and clinical endpoint, indicating that public health goals should dictate key vaccine performance criteria.

Overall, our findings support the need for well-defined target product profiles for long duration vaccines linking priority use cases of where, how, and to whom to deploy new malaria vaccines, to maximize public health impact.

## Introduction

As of 2024, the World Health Organization (WHO) has recommended two pre-erythrocytic vaccines (PEVs), RTS,S/AS01 and R21/Matrix M, for global use against *Plasmodium falciparum (Pf)* malaria in pediatric populations.(1) It is anticipated that vaccine delivery will be aligned with existing routine childhood immunization platforms in perennial settings, or as seasonal mass vaccination before peak transmission among children in areas with moderate to high malaria transmission.(1,2) RTS,S (3) and R21 (4) both act at the pre-erythrocytic stage by targeting the sporozoite surface antigen of the *Plasmodium falciparum* parasite to prevent infection. The groundbreaking WHO recommendation for RTS,S followed three extensive studies confirming the safety and efficacy of the vaccine. These include the RTS,S Phase 3 trials,(5–7) an implementation program for vaccination of children aged five to nine months via the expanded program of immunization (EPI) with a fourth dose at 27 months,(8) and an implementation study of seasonal use case of RTS,S deployed in combination with, or as an alternative to, seasonal malaria chemoprevention (SMC) with sulfadoxine-pyrimethamine and amodiaquine (SPAQ).(9) Most recently, promising evidence from Phase 2 and 3 clinical trials for R21 for both seasonal and perennial use cases reports protective efficacy of more than 75% over 12 months (comparable to RTS,S protective efficacy six months after administration)(10), with further analysis of the follow-up results pending.(11,12) In the short-term, these are likely to be the only malaria vaccines in use, with the choice of implementation strategy predominantly depending on their supply and operational system factors.

Besides RTS,S and R21, other vaccine candidates are in pre-clinical or clinical trial stages, such as the whole sporozoite vaccine *Pf*SPZ(13) and the blood-stage protein vaccine RH5 VLP.(14) There is also a renewed interest and investment in developing novel malaria vaccines, including mRNA vaccines.(15) Development of these new vaccine candidates comes at a crucial time when global progress in the malaria response has stalled.(2,16) Due to drug-resistant parasites, insecticide-resistant mosquito’s, funding needs, climate change and other factors, many African countries are off-track to meet the 2016−2030 Global Technical Strategy (GTS) targets to reduce global malaria incidence and mortality rates by at least 90% by 2030 over 2015 levels.(2) Consequently, the WHO and partners have called for revitalized efforts and the use of new tools to maintain the substantial gains witnessed in previous years and accelerate progress towards malaria elimination. (2) Incorporating a vaccine into the existing and diverse malaria toolbox of interventions is a major milestone that could aid in achieving these targets and increase the proportion of children covered by any intervention.(17) Therefore, there is a need to optimize current vaccine implementation using existing delivery strategies and to understand the preferred vaccine properties, such as efficacy, duration of protection and dosage intervals of new and improved vaccines on their own, as well as alongside other novel interventions for malaria prevention and control.(2) Assessing how such vaccine properties are linked to public health benefits and understanding vaccine performance early in clinical development, including the vaccine’s mode of action and immunogenicity, is essential to support new vaccines to achieve more significant impact. This will enable stakeholders to make informed investment decisions and streamline candidate selection in the Research and Development (R&D) phase of vaccine development.

In 2022, the WHO issued preferred product characteristics (PPCs) for malaria vaccines, providing updated advice on requirements for new vaccine candidates.(18) Informed by multiple stakeholders and public consultation, three strategic goals were identified, the first of which is to develop malaria vaccines that reduce morbidity and mortality in individuals at risk. While the document did not specify strict modes of action, it is outlined in the strategic goals that vaccines are envisaged to provide immunological protection against clinical and severe malaria targeting pre-erythrocytic or blood-stage antigens. Strict efficacy and duration requirements for burden reduction only vaccines were not explicitly defined, though preferred targets against clinical malaria over 12 months were identified. Of note, the PPC highlights that lower clinical efficacy thresholds can be justified in parallel with longer duration of protection, as well as other key drivers of public health impact, including vaccination coverage.(18)

The PPCs document also identified the role of mathematical transmission modelling to support and guide discussions around vaccine impact and performance characteristics. To date, mathematical modelling groups have provided a range of quantitative analyses to support thinking and policy decisions on malaria vaccines. Several studies have used results from RTS,S clinical trials to inform detailed models of malaria transmission and intervention dynamics, predicting the likely population-level health impact and cost-effectiveness of such vaccines. These studies examined vaccines deployed alone or in combination with other malaria interventions,(7) as part of EPI for infants and children,(5) mass vaccination(19), or seasonal use.(7) Other modelling studies have explored target efficacy profiles and decay properties of vaccines for mass vaccination with expanded age groups to support significant prevalence reduction,(20) vaccines for transmission-blocking,(21) or for improved childhood vaccines for EPI use.(22) However, there is still limited evidence from modelling studies of how improving pre-erythrocytic vaccine performance properties and optimizing vaccine deployment could increase public health impact. This includes understanding the impact of vaccine delivery, where boosters are given every other year rather than yearly, and how implementation factors drive vaccine impact.

In this study, we link the full range of vaccine properties, deployment schedules and vaccination coverage to different health outcomes using a detailed simulation model of malaria transmission and vaccines. Firstly, we identify the impact of improving the initial efficacy and duration of vaccine protection on different clinical outcomes, such as averting uncomplicated and severe childhood malaria. Secondly, we investigate how vaccine impact differs by delivery strategy or how impact is driven by system factors such as coverage. More specifically, we focus on understanding the public health impact of implementing improved PEVs, with duration of protection longer than existing vaccines (such as RTS,S), delivered via routine immunization or mass vaccination campaigns followed by annual boosters for five years. We assess vaccine impact by predicting the reduction in infection prevalence, and incidence of clinical and severe cases achieved over the 12 months following the final annual booster in the fifth year. To explore the potential for multi-year vaccine impact, we evaluate the extended protection in the year following primary vaccination for children who do not receive any booster. Through simulation and sensitivity analysis, we provide a quantitative understanding of the trade-offs between vaccine performance and implementation impact-drivers across different endemicities and delivery schedules.

The novel insights from our modelling around the impact of improved PEVs can support the optimization of new malaria vaccine development. Moreover, our findings support improved understanding of current PEVs and their population impact, as well as our understanding of the potential public health benefits of deploying improved PEVs.

## Materials and Methods

### Malaria transmission model

Model simulations were performed using a validated stochastic, individual-based model of malaria transmission in humans, linked to a deterministic model of malaria in mosquitoes, known as OpenMalaria.(23–27) The model is fully open access, has previously been described in (27), and the details regarding our current application with this model are summarized in *S1 Text*. OpenMalaria facilitates impact predictions for a wide range of interventions that target different stages in the parasite life cycle, including bed nets, chemoprevention, and vaccines. Intervention impact can be assessed for various health outcomes, including infection prevalence, uncomplicated and severe malaria cases and deaths. Malaria vaccine strategies have previously been simulated using this model, informed by and calibrated against estimates for RTS,S vaccine’s protective efficacy from clinical trial data(28) and a range of other vaccine trial and implementation data across varying transmission settings.(5,23,28) As outlined below, we simulated a range of vaccine properties, namely, potential efficacies and durations of protection for improved PEVs for different endemicities, seasonal profiles and deployment strategies.

### Simulated model scenarios, settings, and intervention dynamics

The scenarios modelled in this study include a range of vaccine properties of probable values for initial efficacy and half-life duration for PEVs (Table 1). The range of settings represent different archetypal transmission profiles (short season, long season and constant transmission), prevalence levels and intervention coverage levels (reflecting access and uptake). The simulated vaccine deployment schedules encompass vaccine dosage either through EPI or through yearly mass vaccination, co-administration with or without curative malaria treatment and probabilities for accessing antimalarials. These scenarios were developed and refined based on estimates from modelling studies and stakeholder engagement. We further informed the scenarios given our *in-silico* vaccine dose efficacy validation exercise, which used data from a clinical trial on seasonal vaccine use(9) (*S1 Text*). By simulating these scenarios with wide-ranging parameter values, we captured an extensive spectrum of epidemiological malaria dynamics.

**Table 1.** Simulated model parameters.

| Parameter | Description | Parameter range |
| --- | --- | --- |
| **Intervention characteristics** |  |  |
| Intervention | Pre-erythrocytic malaria vaccine (PEV) |  |
| Intervention cohort | Children aged at least five months at recruitment until five years of age |  |
| Population vaccinated with primary doses | Coverage of initial vaccine doses (dose 1-3) | 50–100% |
| Population vaccinated with booster doses | Coverage at each annual boosting of those that received the primary doses (doses 4-7) | 80% of primary series doses coverage |
| **Vaccine properties** |  |  |
| Initial efficacy | Maximum efficacy against pre-erythrocytic infection following three-dose primary vaccination before decay | 50–100% |
| Boosting efficacy | Protective efficacy against pre-erythrocytic infection following the booster dose | 50% and 75% |
| Half-life duration | Time until the initial vaccine efficacy reaches 50% of original value | 6–18 months |
| Decay shape | Weibull function, biphasic decay profile | k=0.69(28), rate of decay parameter, L=half-life duration |
| **Setting characteristics** |  |  |
| *Pf*PR<sub>2-10</sub><sup>#</sup> | Baseline parasite prevalence before intervention (diagnostic detectable) | <10–60% |
| Seasonality | Short season (4-month profile), long season (6-month profile) and constant transmission | 70% of total cases occurring within four or six months, or constant transmission all year round |
| Mosquito species | *Anopheles gambiae* |  |
| **Health system characteristics** |  |  |
| Access to care | Probability of accessing effective curative malaria treatment within 14-days of symptomatic malaria | Default (30%), high (70%) |
| Diagnostic | Rapid diagnostic tests (RDTs) | RDT sensitivity and specificity is 95.0% and 94.2%, respectively |
| **Other interventions** |  |  |
| Co-administration | Blood-stage antimalarial clearance drug modelled as complete blood-stage parasite clearance within a five-day timestep following administration |  |

<sup>#</sup>*Pf*PR<sub>2-10</sub>: *Plasmodium falciparum* parasite rate in 2- to 10-year-olds

#### Vaccine properties

The initial vaccine efficacy against the pre-erythrocytic stage is assumed to reach its maximum level following the primary series. For RTS,S and R21, the primary series includes three vaccine doses, but for new vaccines the primary series may involve fewer doses. In our current study, the vaccine-induced efficacy is assumed to be negligible before administering the final dose in a primary series. Booster doses are assumed to restore the waning vaccine efficacy to levels between 50% and 75% lower than the initial maximum efficacy reached, based on reported values from previous studies(28,29) and our validation results (see next section and *S1 Text*). The decay in protective efficacy over time is assumed to follow a Weibull function with a biphasic shape parameter *k*=0.69, with a rapid decay in the initial months, which is identical both after the initial dose as well as the booster doses as estimated previously.(28) Additionally, in a field trial in seasonal settings, the RTS,S protective efficacy was shown to decline over a three-year trial period, with more rapid decline in the initial six months.(9,10) A two year extension of the same trial also showed sustained protection following additional annual booster doses given before peak transmission until children reached five years of age.(30)

#### Vaccine booster efficacy validation

Using a Bayesian optimization approach (*S1 Text*), we used results from a recently completed Phase three 3 clinical trial for seasonal vaccination with RTS,S in Mali and Burkina Faso(9) to validate the properties of a seasonal vaccine booster. More explicitly, this meant better understanding how the vaccine efficacy following the boosters contrasted against data from the pivotal RTS,S clinical trial, conducted in seven African countries.(6,28) This comparison informed model assumptions around efficacy estimates of the first booster dose given 12 months after the three-dose primary series, rather than 18 months after, as implemented in the original RTS,S trial.(6,28) The initial vaccine induced efficacy following the primary series, 91.1% [95% CI 74.5–99.7%], was used as a model input and this efficacy was separately reproduced *in silico* using the OpenMalaria model to match the trial results (*S1 Text*). The booster doses, given 12 and 24 months following the primary series, were assumed to partially restore waning efficacy, although it remains unclear by how much and for how long protection is extended. We used our Bayesian approach, which utilizes a Gaussian process regression model as the objective function(31), to find the optimal parameter values for booster efficacies for the different trial arms, that minimize of the residual sum of squares between the observed data and modelled outputs.

#### SMC with SPAQ half-life validation

Similarly, using the Bayesian optimization approach described and the results from the same Phase 3 clinical trial in Mali and Burkina Faso(9), we sought to calibrate the preventive half-life duration of seasonal chemoprevention using SPAQ. In the control arm, SPAQ was deployed to a cohort of children alone then in another arm in combination with seasonal vaccination. We incorporated past estimates of initial efficacy following dosing with SPAQ as inputs for our model.(32) We assumed that SPAQ acts by first clearing all blood stage infections, followed by preventive action represented by a Weibull decay function with shape parameter k = 5.40.(32)

#### Vaccine deployment

As described above, we defined primary vaccination as receiving the first series of doses (for example, up to three doses) and full vaccination as receiving the primary series of doses and annual boosters up to age five. In this study, all vaccinations are delivered through two approaches to allow comparison between deployment schedules. In the first approach, which we refer to as hybrid vaccination, we deploy the three-dose primary series as part of the age-based immunization schedule. The initial vaccine doses are given continuously during the intervention period to children aged five, seven and a half, and nine months. In the second approach, which we refer to as mass vaccination, children aged between five and 17 months receive the three-dose primary series through a mass campaign, timed so that the third dose is given one month before the transmission season’s peak. For both approaches, additional annual boosters are deployed over four years to the same children up to five years of age, one month prior to peak malaria transmission in seasonal settings (Fig 1). In settings where the transmission is constant throughout the year, we follow the same delivery schedules for the primary series with the additional booster doses given through yearly mass campaigns (*Fig A in S1 Text*). The dropout rate is assumed to be 20% for all booster doses compared to the primary series. Vaccine doses are deployed singly or co-administered with a highly efficacious antimalarial treatment, modelled as blood-stage parasite clearance over five days to represent treatment with artemether-lumefantrine.

**Fig 1:**
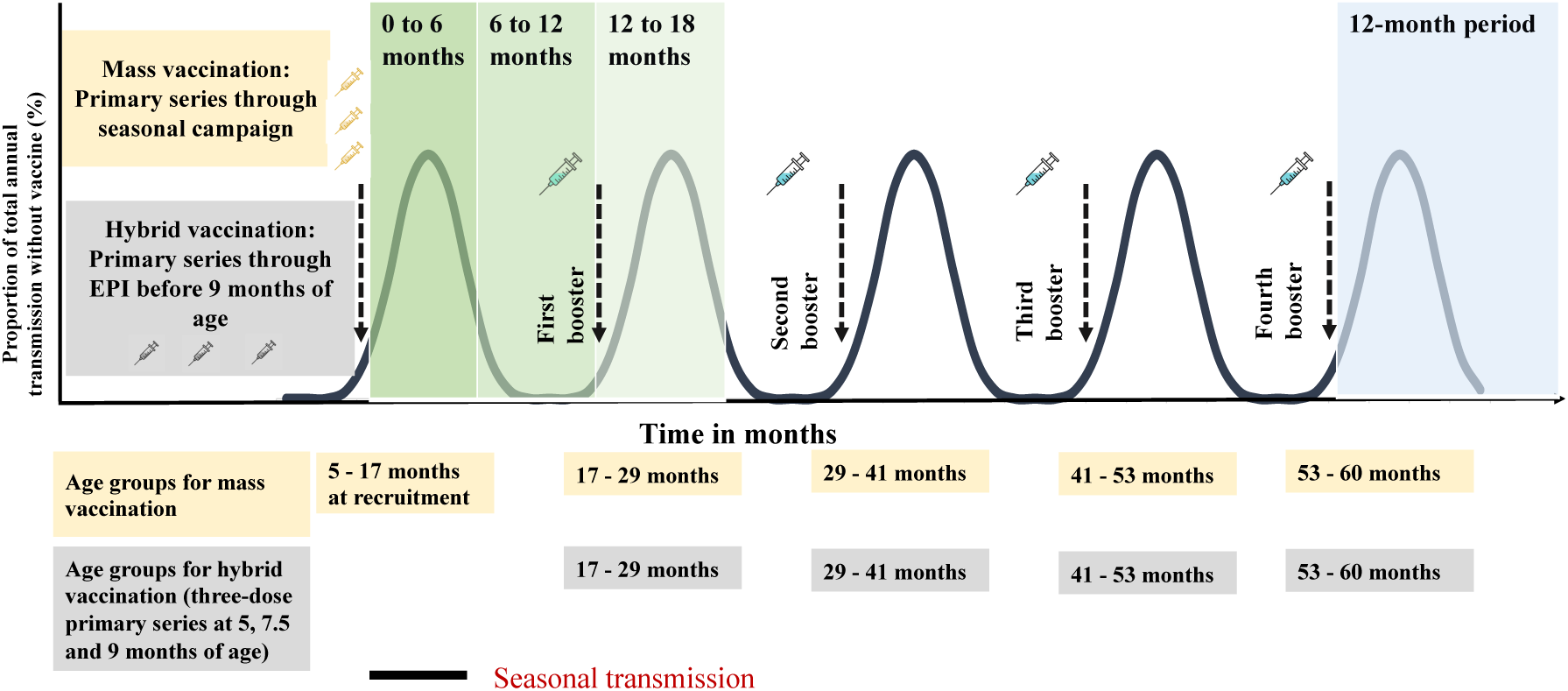
Schematic illustration of the simulated vaccine deployment schedules for a five-year vaccination program shown for a seasonal setting. Illustration shows the five-year vaccination program with a primary series dosage, the timing of the annual boosters and at the bottom of the figure the age groups at each dose for both vaccine delivery schedules, hybrid vaccination (grey boxes) and mass vaccination (yellow boxes). For the mass vaccination schedule the primary series and annual booster doses are deployed before the peak transmission season. For the hybrid vaccination schedule doses for the primary series are deployed continuously as part of an age-based immunization schedule while the annual boosters are also deployed before the peak transmission season. The zero to 18 months period (green shaded areas) shows when the multi-seasonal or multi-year vaccine impact is evaluated by comparing a cohort of children who received the primary series doses and the first booster (dose four) and those who only received the primary series doses. The 12-month period (grey shaded area) follows the final annual booster dose (dose seven) in the fifth year where the public health vaccine impact is evaluated for children who received the primary series followed by annual boosters.

### Endpoints to assess vaccine impact

Three public health outcomes were evaluated, including the relative reduction in infection prevalence, incidence of clinical cases, and incidence of severe cases, all compared to a no-intervention counterfactual. All three health outcomes were evaluated for two target age groups and follow-up periods. First, the vaccine impact was evaluated in children aged six years and below, 12 months following the final annual booster dose (dose seven) in the fifth year. We estimated vaccine impact across all the simulated scenarios for the two deployment schedules (Fig 1). Second, we evaluated the multi-year vaccine impact in the two years following the three-dose primary series by comparing a cohort of children who received only the primary series with those who received the primary series plus one booster dose (dose four). For the multi-year impact, the 24-month follow-up period was divided into six-month intervals. This multi-year impact is intended to assess a vaccine’s extended protection in the first and second years if children do not receive booster doses, particularly in seasonal settings.

### Statistical and global sensitivity analyses to evaluate vaccine impact

Using our individual-based stochastic malaria transmission model, we simulated experiments matching the scenarios described above. For the vaccine properties (initial efficacy and half-life duration) and coverage, we generated a Latin hypercube of 1000 samples, and for each combination simulated outcomes for five replicates. We used heteroskedastic Gaussian process regression (hetgp package in R(33)) for each scenario to fit a model emulator to our database of model simulations. This emulator could capture the relationship between key vaccine performance properties and other factors, such as coverage or access to treatment, as regression inputs, and health outcomes, as outputs.(34) Exploring the entire parameter space for the different combinations of vaccine properties, deployment schedules, endemicities, and seasonality profiles requires a large number of simulations, which is computationally intensive. At low computational cost, emulators captured the relationships between vaccine properties and system factors, as well as the predicted health outcomes.(20) These emulators were then used to predict the vaccine’s impact over the different follow-up periods and target age groups. We evaluated emulator performance by testing 10% or the total simulations against 90% used in the training set.

To identify the most important drivers of vaccine impact for different settings, health outcomes, and follow-up periods, we conducted a global sensitivity analysis of our Gaussian process regression model results using the Sobol method(35) and reported total effect indices. To calculate the relative contribution of each property, the total-order effect indices were normalised. The sensitivity analysis measured the extent to which a small change in an intervention’s key performance property corresponded to a change in its impact. For example, an increase in the initial maximum vaccine efficacy from 50% to 90% may lead to a larger change in the achievable clinical incidence reduction than a six-month increase in the vaccine’s duration of protection. All analyses were conducted in R-software (version 4.1.0).(36)

## Results

### Validation of vaccine booster efficacy using clinical trial data

Using the earlier described Bayesian optimization approach(31), we could determine booster efficacy estimates from the clinical trial arm where only the RTS,S vaccine was deployed and in the arm when combined with SMC. By incorporating past estimates of the vaccine’s initial efficacy following the three-dose primary series of 91.1% and the half-life duration of 7.32 months as model inputs,(28) we were able to approximate observed data from the trial to match our simulated modelling outcomes in both countries. Assuming a biphasic decay in vaccine efficacy and our Bayesian optimization approach (*S1 Text*), we estimated a best fit to incidence data for the vaccine’s maximum efficacy after the first booster (dose 4 given 12 months after dose 3) of between 80.50% to 88.00% in the vaccine only arm and 80.50% to 85.40% in the combination arm in Burkina Faso (high prevalence, seasonal transmission) and Mali (moderate prevalence, highly seasonal transmission). Following the second booster (dose 5, given 12 months after dose 4), the maximum boosting efficacy was reduced to between 65.95% to 80.00% in the vaccine only arm and 76.48% to 80.00% in the combination arm in these countries. Fig 2 shows the monthly clinical incidence throughout the study period, reflecting the known estimates for initial efficacy achieved following primary vaccination and the estimated assumptions for the efficacy achieved following the two booster doses (doses four and five) administered prior to the peak malaria season in Burkina Faso and Mali that best fit the clinical trial data.

**Fig 2:**
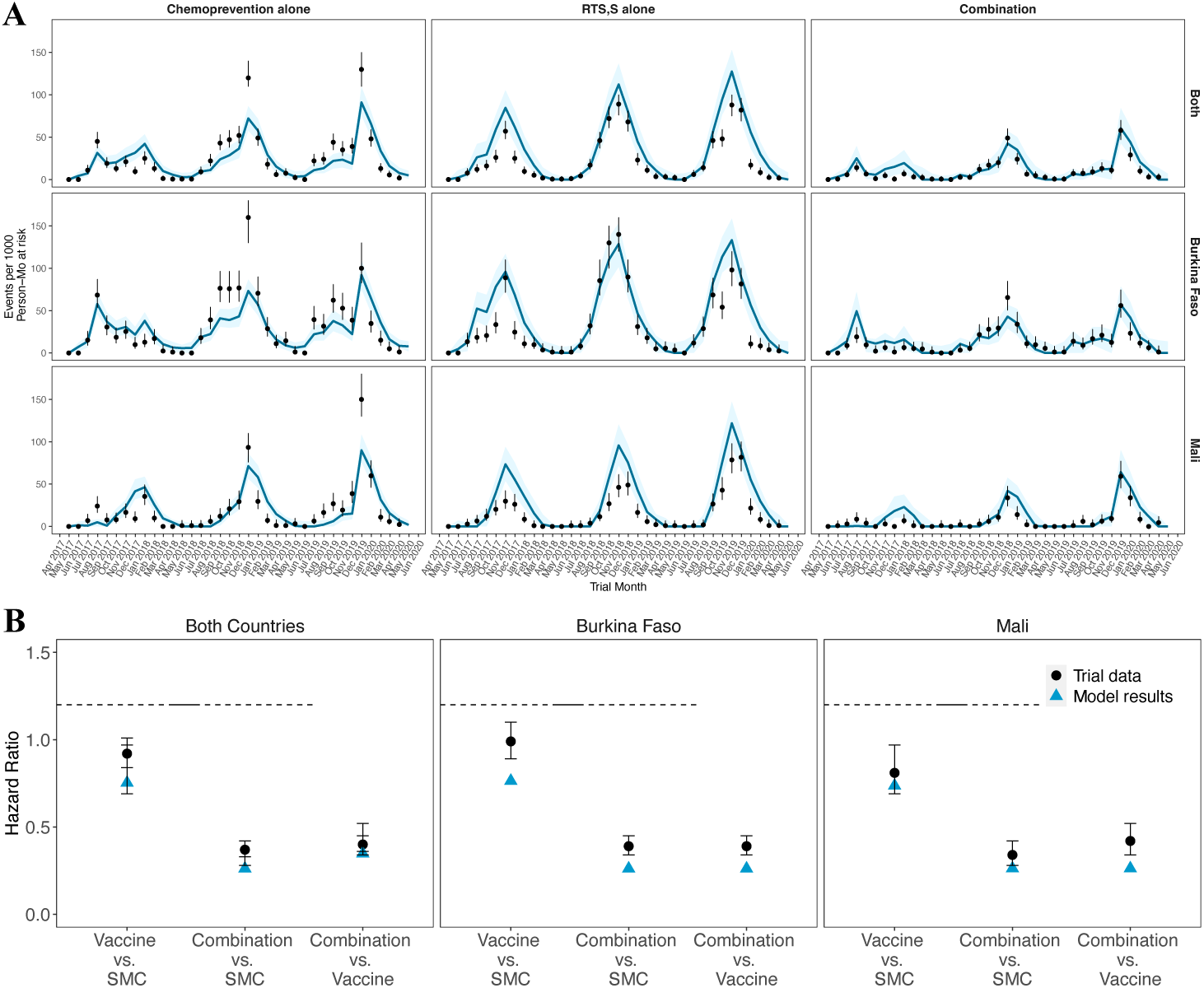
Monthly clinical incidence and hazard ratios for the clinical trial data (black dots) compared with model simulations (blue lines/triangles) using the best fit assumption for the efficacy of annual booster doses four and five and SPAQ preventive half-life duration. A) Best fit for Burkina Faso (*Pf*PR_6-12_ is 50−60% and seasonal transmission), Mali (*Pf*PR_6-12_ is 20−30% and highly seasonal transmission) and both countries combined shown for the three trial arms, B) Hazard ratios between the trial arms for both countries separately and combined. The black dots shown with 95% confidence intervals represent the trial field data and the blue lines/triangles illustrate the modelled output from the simulations with the shaded region showing the confidence intervals averaged over 100 seeds. In this figure, the parameters were optimized for the arms where chemoprevention or vaccination were given alone and used to predict the model results for the arm where the vaccine and chemoprevention were combined.

The vaccine boosting efficacy estimates varied between the two countries and also within each country, exhibiting broad error margins of uncertainty. The estimates ranged from 5% to 30% lower than the initial efficacy against infection reached after primary vaccination (Fig 2 and *Fig C in S1 Text*). These differences could be attributed to reported variations in malaria epidemiology between the two countries, as measured in children aged six to 12 years old at the end of the peak transmission seasons during the trials, but also likely due to differential profiles of exposure and acquisition of immunity in the non-vaccine SMC arm (larger drop in 4^th^ dose efficacy in the higher transmission site).(9) Thus understanding of country- or endemicity-specific evaluation of vaccine performance in addition to global or archetypical estimates of vaccine efficacy and duration is important. Hazard ratios calculated from both countries were also shown to fit the clinical trial data for both countries across all the trial years (*Fig 2, Fig D in S1 Text)*.

### Validation of SP-AQ preventive half-life using clinical trial data

We replicated the optimization approach for the chemoprevention only arm where SPAQ was deployed for four monthly cycles each year of the trial and in the arm where chemoprevention was combined with seasonal vaccination. We estimated the optimal range for the half-life duration of protection to be between 20 days and 27 days in both arms in both Burkina Faso and Mali. These estimates were lower than what has been reported previously from clinical studies(37) and through modelling.(32,38) We were also able to approximate the decay shape parameter which matched our simulated modelling outcomes in both countries. This ranged between 2.55 and 4.30 in Burkina Faso, and between 4.30 and 5.30 in Mali. Fig 2 shows the monthly clinical incidence rate and cumulative hazard estimates throughout the study period, reflecting the best fit assumptions for the half-life following administration of SPAQ administered prior to the peak malaria season in Burkina Faso and Mali. Results for the trial arm where RTS,S was deployed in combination with SMC are also shown in Fig 2, with additional results on best fit in *Fig C in S1 Text*.

### Public health impact of improved PEVs on Plasmodium falciparum malaria burden

Modelling results indicate that implementing an improved PEV, targeting children aged over five months at recruitment, is expected to yield substantial impact on reducing infection prevalence, followed by decreases in clinical incidence, and then severe disease, when assessed among children under six years of age (Fig 3). As described above, vaccine impact is evaluated in the 12 months following the final annual booster dose in the fifth year. Across all modelled scenarios, our findings consistently confirm that co-administering a PEV with a blood-stage parasite clearance drug leads to a substantially greater reduction in disease burden compared to vaccination alone (*Fig E in S1 Text*). Subsequent sections present results for scenarios where each vaccine dose was co-administered with a blood-stage clearance drug.

**Fig 3.**
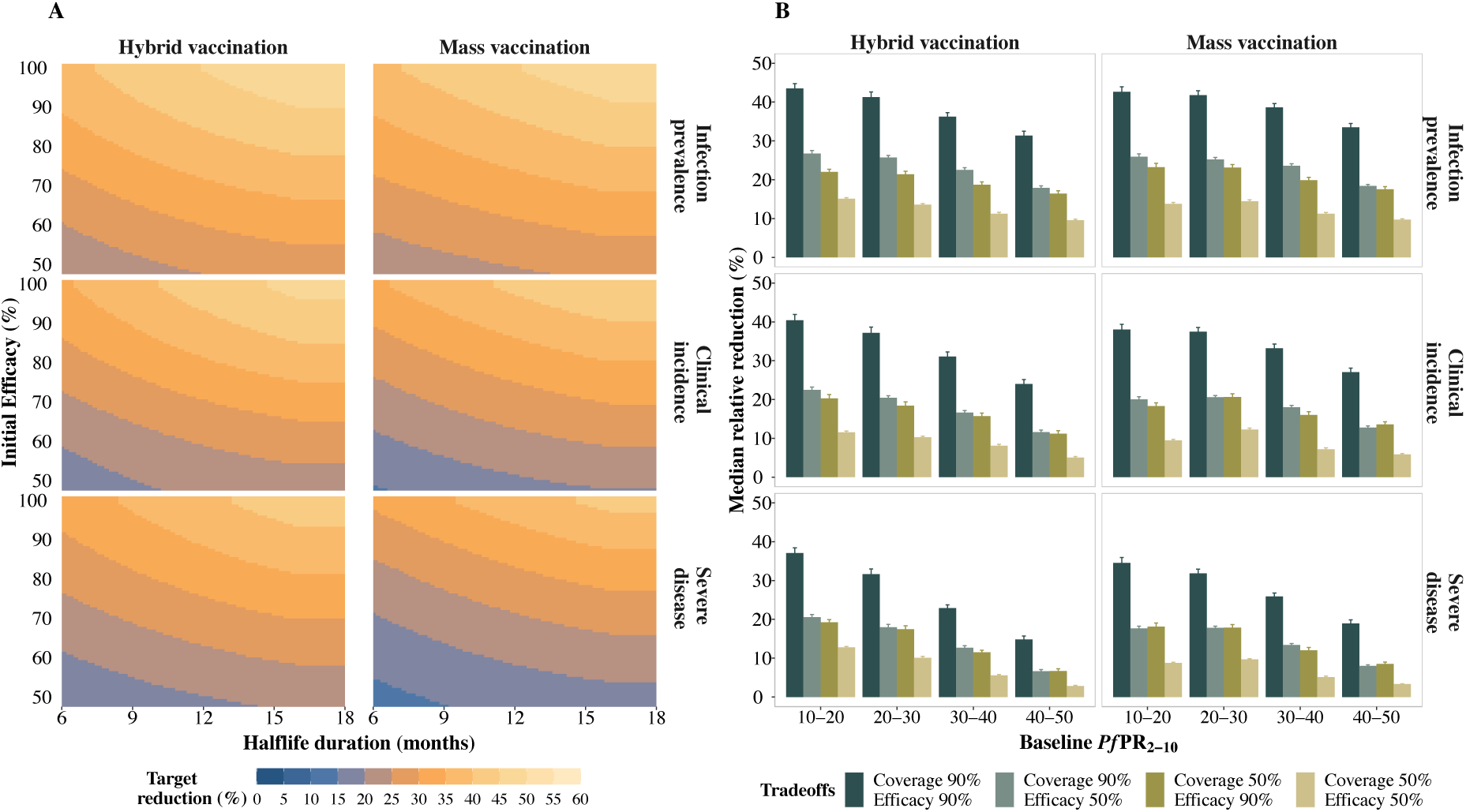
Predicted relative burden reduction in the 12-month period following the final annual booster dose, compared with a no-intervention counterfactual. A) Target reduction (%) in infection prevalence (top row), clinical incidence (middle row), and severe disease (bottom row) illustrating trade-offs between initial efficacy and half-life duration of protection in settings where baseline *Pf*PR_2-10_ ranged between 10% and 20%. The initial efficacy ranged from 50% to 100%, half-life duration from six to 18 months, and assuming a primary series vaccination coverage of 90%. B) Median (interquartile range (IQR)) relative reduction in infection prevalence (top row), clinical incidence (middle row), and severe disease (bottom row), considering varying levels of baseline *Pf*PR_2-10_, coverage, and initial efficacy for a long duration vaccine with a half-life duration between 12 and 18 months. Results are shown for a PEV co-administered with a blood stage clearance drug, for a four-month short seasonality profile, for both deployment schedules, in settings with a 30% probability of accessing curative treatment within 14 days of symptom onset.

The most notable relative reduction in disease burden occurred in areas with low and low-moderate transmission (*Pf*PR_2-10_ <30%) and decreased with increasing transmission (Fig 3b). Vaccine impact defined as the relative reduction in burden 12 months after the final annual booster, increased with improved vaccine performance and higher vaccination coverage (Fig 3a, 3b). Improving the initial efficacy by, for instance, increasing it from 50% to 90% or by increasing vaccination coverage from 50% to 90%, resulted in almost a twofold increase in impact. Vaccines with extended half-life durations could offer protection for multiple years, while booster doses strengthen this protection, albeit contingent on dropout rates. From our trade-off analysis, our results show that we still need high initial efficacy if the half-life duration is less than 12 months. Increasing this half-life to between 12 and 18 months or reaching more children can allow us to achieve the same impact with lower levels of vaccine efficacy (Fig 3, *Fig H in S1 Text*).

Across all contexts, burden reduction varied only slightly between the two delivery schedules (Fig 3). In highly seasonal settings, the hybrid vaccination schedule demonstrated a marginally better potential when compared to settings with longer seasons where the mass vaccination schedule showed a higher impact (*Fig F in S1 Text*). This preference stems from the fact that when vaccines are administered through mass campaigns, they simultaneously provide maximum protection to more children and this, in turn, leads to a greater impact on the transmission dynamics and, as such, to less infections. Conversely, administering the primary series continuously through a hybrid schedule could provide more protection, especially during short peak seasonal transmission. We observed similar findings in settings where transmission is assumed to be constant throughout the year. Burden reduction was slightly more pronounced when all vaccine doses were deployed through yearly mass campaigns, as opposed to hybrid vaccination (*Fig F in S1 Text*). We also found that, in all settings, relative burden reduction was substantially lower if health systems were stronger, particularly where higher levels of access to curative malaria treatment for a clinical case were available (*Fig G in S1 Text*).

### Factors influencing vaccine impact following the five-year vaccination program

Reaching more children with a vaccine leads to greater impact. From our sensitivity analysis, the most important driver of impact is the proportion of children reached with the primary series doses, or vaccination coverage, followed by the vaccine’s initial efficacy and half-life duration. This is especially the case for infection-related endpoints and on a smaller scale, when vaccine administration is through hybrid vaccination compared to the mass vaccination schedule (Fig 4).

**Fig 4:**
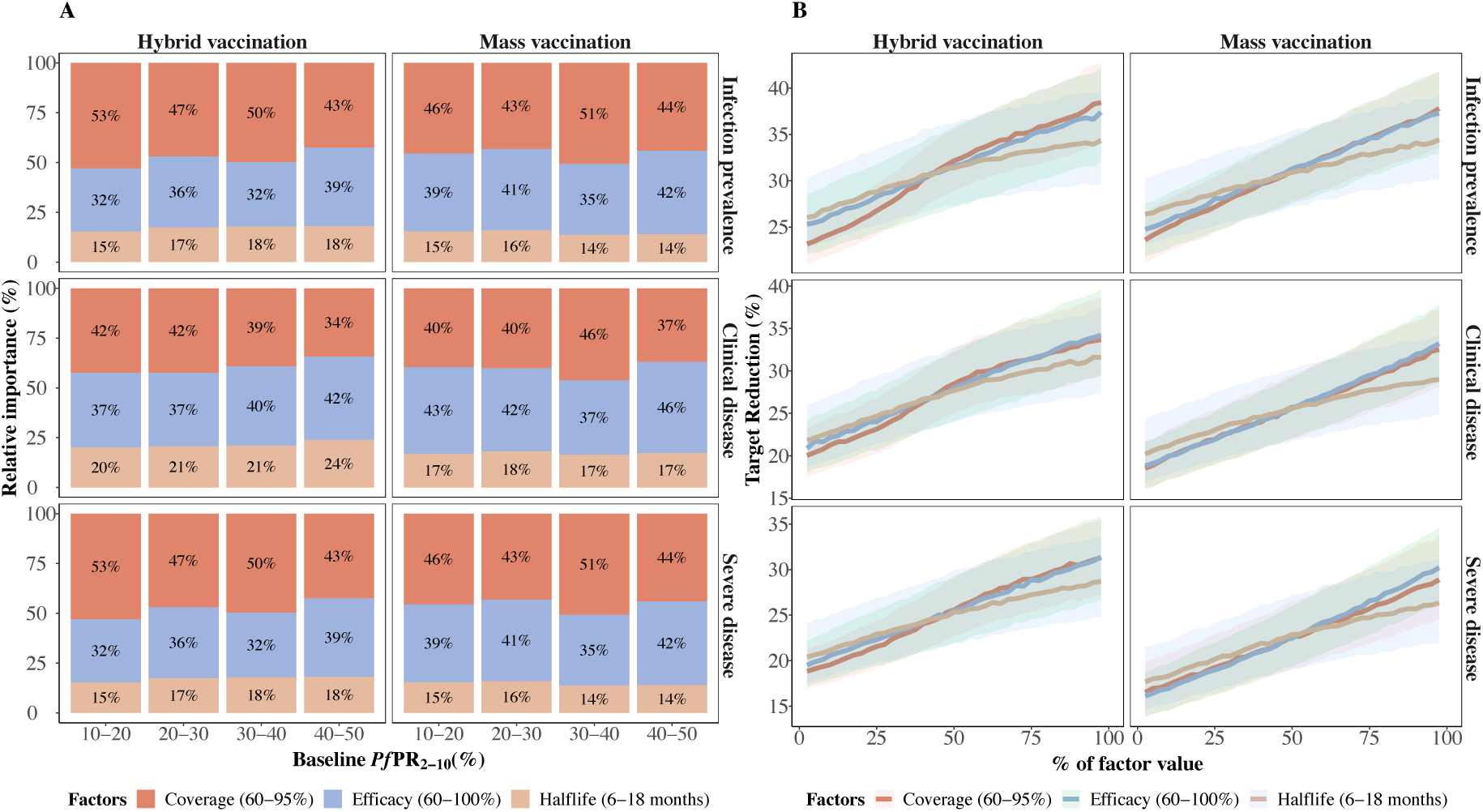
Factors influencing vaccine impact on predicted burden reduction for the 12 months period following the final annual booster dose compared with a no-intervention counterfactual. A) Bars indicate the total Sobol effect indices, which explain the variance in predictions of relative reduction in infection prevalence (top row), clinical incidence (middle row), and severe disease (bottom row). These indices can be interpreted as the proportion of variation in the outcome attributed to changes in each variable. Results are shown across various baseline *Pf*PR_2-10_ values and span different parameter ranges for initial efficacy (70% to 100%), half-life duration (six to18 months), and vaccination coverage (60% to 90%). B) The influence of the impact-driving factors on predicted reduction in infection prevalence (top row), clinical incidence (middle row), and severe disease (bottom row) for settings where *Pf*PR_2-10_ lies between 20% and 30%. The different lines and shaded areas depict the median and interquartile range (IQR) of proportional contribution, as estimated through global sensitivity analysis over the variable parameter ranges for initial efficacy (60% to 100%), half-life duration (six to 18 months) and vaccination coverage of (60% to 95%). Results are shown for a PEV co-administered with a blood stage clearance drug, for a 4-month short seasonality profile, for both deployment schedules, in settings with a 30% probability of accessing curative treatment within 14 days of symptom onset.

We assessed outcome metrics over a range of baseline prevalence levels and seasonality patterns. While the relative reduction in disease burden differed between the modelled settings, our key findings underscore the crucial role played by both the initial efficacy against infection and the half-life duration of protection. We found that the relative contribution of each vaccine property to the overall impact depends on the clinical endpoint of interest, the seasonality patterns, the timing and length of the evaluation period, and whether the vaccine was co-administered with a blood-stage clearance drug (Fig 4 and *Fig I, J in S1 Text*). For instance, when endpoints are evaluated closer to the biological time of action, such as the 12 months following the final annual booster, burden reduction is primarily driven by the initial efficacy, as shown in Fig 4. However, by enhancing the initial efficacy to >90% and achieving an 80-95% coverage with primary doses, the half-life duration becomes the predominant driver for burden reduction following the final annual booster, which in some instances accounts for over 60% of the total impact. While our findings regarding the significance of initial efficacy and half-life duration hold consistent across different delivery schedules, our results suggest higher requirements for vaccine performance when existing infections are not pre-cleared during vaccination. The vaccine properties drive most of the impact in these cases, particularly for severe clinical outcomes (*Fig I in S1 Text*).

### Multi-year vaccine impact in the two years following primary vaccination

To estimate the extended protection in the second year following primary vaccination, we evaluated impact in the 24-month period by following children who did not receive booster doses (Fig 5). Multi-year impact was evaluated by comparing children who received only the primary series doses in the first year to those who received the primary series doses plus one booster (fourth dose) in the second year (Fig 5, *Fig M in S1 Text*). Without the booster, a modeled PEV with a half-life between six and 12 months still provided extended protection beyond the first year following primary vaccination, covering part of the subsequent year’s second season (Fig 5). Notably, when the half-life duration of protection was longer than 12 months, a substantially higher burden reduction was predicted in the second year. This was due to the vaccine’s protection extending to encompass the entirety of the second season, although at reduced efficacy. This phenomenon was particularly apparent in settings with pronounced seasonality, characterized by shorter periods of higher transmission, suggesting that the vaccine could be classed as multi-seasonal or multi-year (Fig 5). With yearly boosters given through mass vaccination in seasonal settings, our results show a 60-80% incidence reduction each year (*Fig M in S1 Text*), consistent with seasonal RTS,S and R21 trials in settings where *Pf*PR_2-10_ <30%. For both cohorts, our findings show a higher impact during the second year when the primary doses were deployed through the mass vaccination schedule before the first year’s peak season, in comparison to hybrid vaccination. This could be attributed to the mass deployment of primary series doses to more children simultaneously in the first year, preventing a higher proportion of infections even when booster doses were not given (Fig 5, *Fig L in S1 Text*).

**Fig 5:**
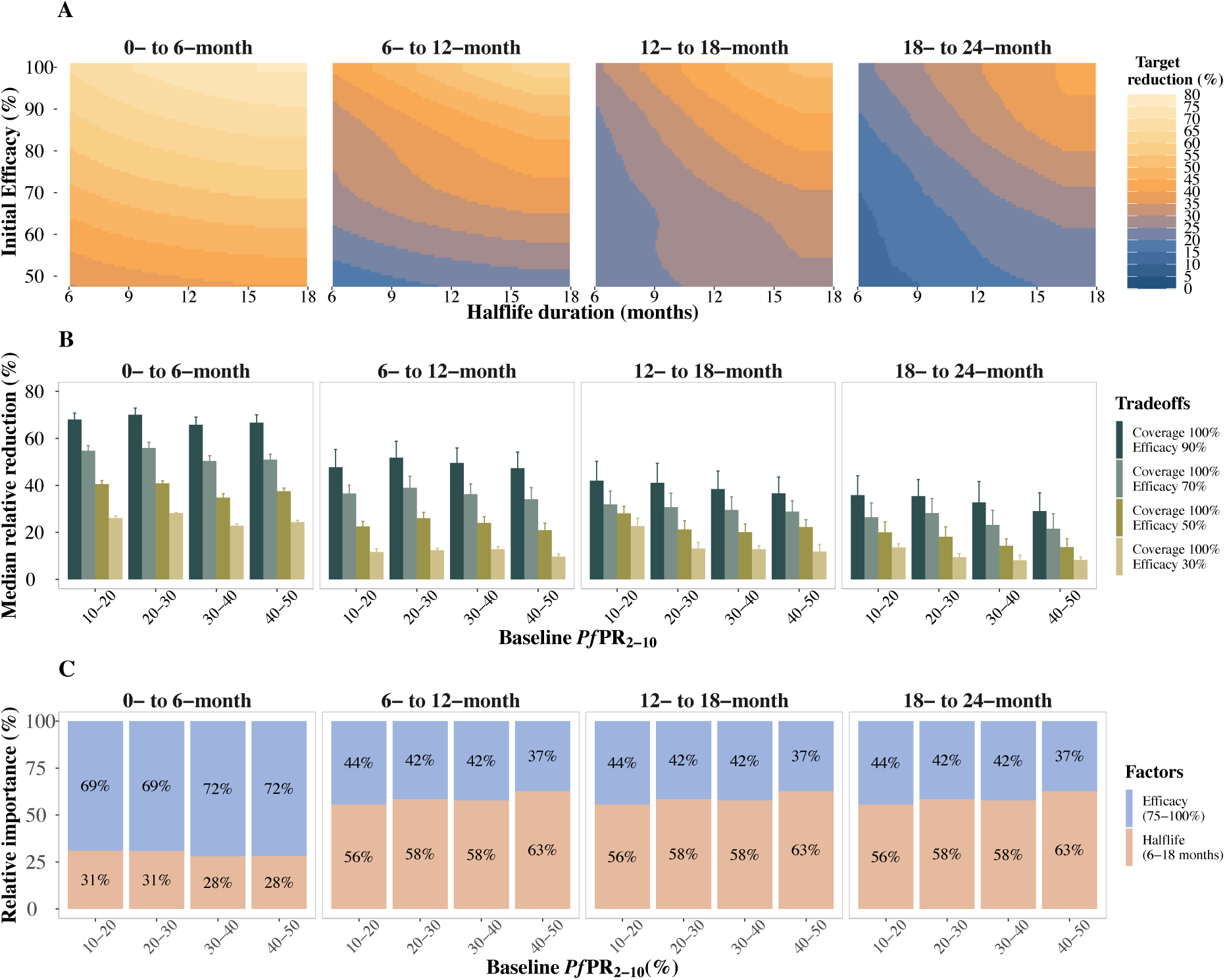
Predicted relative reduction and impact-drivers on clinical incidence in the 24 months following primary vaccination for children who did not receive a booster, compared to a no-intervention counterfactual. A) Trade-offs between initial efficacy and half-life duration of protection for clinical incidence reduction in settings where baseline *Pf*PR_2-10_ ranged between 10% and 20%, initial efficacy ranged from 50% to 100%, and half-life duration from six to 18 months, assuming a primary series vaccination coverage of 100%. B) Median (interquartile range (IQR)) relative reduction in clinical incidence for different levels of *Pf*PR_2-10_ and initial efficacy, for a long duration vaccine with a half-life between 12 and 18 months and primary series vaccination coverage of 100%. C) Bars indicate the total Sobol effect indices, which explain the variance in predictions of relative reduction in clinical incidence. These indices can be interpreted as the proportion of variation in the outcome attributed to changes in each variable. Results are shown over different parameter ranges for initial efficacy (75% to 100%) and half-life duration (six to 18 months), where vaccination coverage was fixed at 100%. Results are shown for a PEV co-administered with a blood stage clearance drug, for the zero- to six-, six- to 12-, 12- to 18- and 18- to 24-month periods following primary vaccination, for a four-month short seasonality profile, for the mass vaccination deployment schedule, in settings with a 30% likelihood of accessing curative treatment within 14 days of symptom onset.

Compared to the factors driving vaccine impact after five years of vaccination with boosters in previous sections, our analysis of multi-year impact confirms the vital role played by both the initial efficacy against infection and the half-life duration of protection (Fig 5c). Vaccinating all children in the cohort with the primary doses, we found that the importance of each vaccine property in driving impact varied by the delivery schedule and follow-up period. The impact of a multi-year PEV with an initial efficacy >75% (comparable to existing vaccines like RTS,S) during the 12 to 24 months following primary vaccination is determined by the duration of antibody protection (Fig 5c and *Fig L in S1 Text*). However, while the initial efficacy is a less influential driver for extended protection in the second year, a multi-year vaccine still needs to have a relatively high initial efficacy to achieve adequate burden reduction (Fig 5). Such a vaccine would require a half-life duration longer than existing vaccines (at least 12 months) to achieve a burden reduction in the second season that is at least half of that estimated in the first season (Fig 5a, 5b and *Fig L in S1 Text*). For instance, in both the deployment schedules and without boosters, to achieve a >30% reduction in clinical incidence in the 12- to 18-month period following a >60% reduction during the zero- to six-month period after primary vaccination, a half-life duration of at least 12 months and an initial vaccine efficacy of >70% are both required, if the vaccination coverage is 100%. The initial efficacy drove most of the impact in the six-month period following the mass vaccination schedule (Fig 5c). However, this was not the case for the hybrid vaccination approach, where the main driver of impact was the half-life duration across all the follow-up periods (*Fig L in S1 Text*).

## Discussion

Our modelling results show that PEVs with improved duration of protection and vaccine-induced protective efficacy have the potential for increased benefit in reducing childhood malaria. Moving beyond current CSP-targeting vaccines (i.e., RTS,S), we provide additional evidence on the trade-offs and relationships between vaccine performance and drivers of impact for these improved vaccines. Longer duration PEVs could provide extended protection in the years following primary vaccination, and while efficacy wanes over time, boosting can restore protection. Our results suggest that deploying a PEV with a half-life duration that could be extended by three to five months compared to that of RTS,S may result in sustained impact into the second and third year following primary vaccination. If the initial efficacy for a new vaccine candidate is more than 90%, improving the half-life duration to more than 12 months can achieve a burden reduction of more than half relative to the preceding year’s level, even when a booster is not given in the second year. Moreover, if the duration of protection can be increased further, trade-offs could be made with lower initial efficacy for a similar impact. This means there is a crucial need to reliably measure the duration of protection of vaccines. While it is currently challenging to adequately measure duration before large scale clinical trials, early evidence could be measured in a controlled human malaria infection (CHMI) trial alongside reliable immune correlates of protection.

Recent evidence has shown promising results for the RTS,S vaccine delivered through EPI or seasonal mass campaigns,(8,9,30) with additional encouraging results reported for R21.(11,12). While these might be the only vaccines available for a few years, our findings indicate that use cases for improved PEVs should adapt and capitalize on the benefits of multi-year protection. If a new vaccine can provide multi-year protection, a program can reduce the number of booster doses to be delivered to at least every two years instead of annually. In addition, when implementing multi-year PEVs with a duration of protection greater than 12 months, children missed during annual seasonal vaccination will remain partially protected. If more children are reached with the vaccine, individuals could also benefit from the indirect effects of reduced malaria transmission across the entire population. Developing such a vaccine and combining it with other malaria interventions, such as vector control or chemoprevention, can potentially reduce severe disease burden, save lives, and accelerate elimination efforts.

We parameterized vaccine boosting efficacies and SMC properties in our model using trial results of RTS,S/SMC.(9) Accounting for trial site characteristics we were able to match our modelling results to trial data from both Mali and Burkina Faso. We also reproduced trial findings that showed increased impact following seasonal boosting, similar to our modelling results. These findings emphasize the need to understand how vaccines work in settings with varying transmission dynamics or how vaccines interact with other interventions. In particular, we recognize the importance of clearly defining temporal transmission profiles for vaccines to ensure they are deployed before or during the maximum risk period in seasonal settings for increased impact. We carefully explored the parameterization of the RTS,S vaccine given alone or in combination with SMC in these seasonal settings, thus also validating intervention properties. Similar to the clinical trial findings (9), we found higher impact when combining the PEV with antimalarials. From our modelled scenarios, co-administering a PEV with a blood-stage clearance drug to clear existing infections leads to higher impact than when the vaccine is deployed alone. It has previously been shown that combining vaccines with mass drug administration in the final stages of an elimination program, may enhance the success of interrupting transmission as opposed to single deployments.(19) Where the goal has been predominantly to reduce burden, higher impact was seen when RTS,S was deployed in combination with antimalarials, as shown in a Phase 3 trial for seasonal vaccination plus SMC using SPAQ(9,30) and in a Phase 2 trial combining RTS,S with dihydroartemisinin, piperaquine, or primaquine.(39) Interactions with combination drugs should be investigated and resolved to understand their relative and joined efficacies, as well as to cultivate community uptake of such treatment strategies.

Overall, we show that the impact of PEVs on reducing burden is both more consistent across transmission settings and higher for infection endpoints than clinical and severe outcomes. This indicates that, considering alternative clinical endpoints may allow better evaluation of vaccine efficacy in Phase 2 and 3 clinical trials conducted in diverse settings. For PEVs, an infection endpoint is closer to the mode-of-action of the vaccine and this endpoint can be translated across settings and, furthermore, offers a systematic and unbiased way to compare different malaria vaccines. At present, efficacy endpoints for current vaccines are generally reported against uncomplicated and severe disease. Endpoints from existing trials are difficult to interpret and compare, as they are measured for varying transmission settings, follow-up periods, and underlying age patterns of disease.(6,12,13) Future clinical trials could evaluate all infection endpoints with sensitive diagnostics or appropriate serological monitoring, particularly if vaccines are to be considered as part of the toolkit for achieving malaria elimination. Monitoring the incidence of any malaria infection in a clinical trial sub-cohort will confirm the underlying efficacy of a vaccine against infection Moreover, validating immune correlates to inform the early phases of clinical trials, including CHMIs, will support the evaluation of dosing regimens prior to conducting larger scale, later-stage clinical trials. This will be essential for reducing the time required to evaluate longer duration vaccines.

Our modelling results show that, for all health outcomes examined, impact increased with improvements in vaccine performance and the proportion of children vaccinated, with more pronounced changes to impact for duration of protection improvements. While it may take several years to develop and approve a next-generation malaria vaccine (such as multi-stage vaccines or non PEV), improving the efficacy, duration, or the delivery of PEVs could improve their impact. Furthermore, as presented in this study, modelling new candidates with optimized deployment strategies, such as through decreased doses or timed deployment to increase coverage, or both, will aid in the assessment of the required performance characteristics for their suitability as novel vaccines. Product characteristics which could influence vaccine coverage include its formulation and the number of doses required, as they could impact acceptability and adherence, cold storage requirements and a potential low frequency of adverse effects. All such considerations for a prospective vaccine candidate should be evaluated in parallel with determining their initial efficacy, duration of protection, and the scheduling of booster doses. Based on the results from our trade-offs analysis, we find that a vaccine candidate with a low initial efficacy of 50%, less than for RTS,S, would likely have substantially lower impact, an outcome which is contingent on vaccine durability as well as factors such as health system capacity in the affected community. However, an implementation strategy that seeks to reduce the number of vaccine doses per child at the cost of reducing vaccine efficacy while, at the same time, improving vaccination coverage to allow two rather than three vaccine doses per child during the primary series, may still translate to a public health benefit. This implies that continued efforts to develop vaccine candidates that are less efficacious than RTS,S remain worthwhile, even in the context of the current focus for vaccine development strategies to maintain protectiveness over time.

From our results, reaching children with a vaccine is the most important determinant of vaccine impact, particularly for low efficacy or shorter duration PEVs. This underscores the importance of evaluating and understanding both vaccine properties and operational factors that influence intervention access, delivery, and uptake. Particularly, in-addition to reducing primary series doses, if vaccines are delivered through routine health systems. Improving coverage, for instance, by matching current booster dose timing to routine immunization schedules, fractional dosing, reducing supply and demand gaps, or sub-national targeted vaccination, could improve the expected impact.(22,40) Several challenges, including health system constraints, gaps in communication and engagement between stakeholders, and inadequate training and community sensitization, were identified in the piloting of RTS,S in Ghana, Kenya, and Malawi.(17,41,42) These are postulated to have contributed to inefficiencies in delivery, impeding high vaccine uptake.(43) The timing of the RTS,S fourth dose was also not aligned with existing childhood immunization schedules (the booster was given at 27 months of age whilst the more common measles and meningococcal vaccines are given at 18 months of age). Additionally, the age eligibility criteria did not match the burden in some countries.(17,43) We did not explicitly explore the influence of the efficacy of all doses in the primary series, or of reducing the number of primary doses in our study. However, a competitive edge could be provided by a PEV with fewer doses at a lower cost, with an advantage in supply, or with safety and efficacy demonstrated in school-going children and adults.

Our study has several limitations to consider when interpreting the results. First, we based our model parameterizations for an improved PEV on data and estimates from two clinical trials: a Phase 3 trial of RTS,S that included the likely protective efficacy decay profile after a three-dose primary series,(28) and a trial of seasonal vaccination with RTS,S in two countries(9) to validate model assumptions of boosting efficacy. Estimates surrounding boosting efficacy have yet to be exhaustively validated in the field and are hence uncertain. While our validation exercise captured some of these uncertainties, future modelling studies must describe these ambiguities fully. In particular, it will be critical to onboard new clinical evidence or knowledge around a new vaccine candidate’s decay in protection. Better informed efficacy decay parameter estimates will directly improve model predictions of the potential PEV impact. Second, our findings are influenced by our assumptions on vaccine performance properties, including the selected parameter ranges for initial efficacy and half-life duration of protection. We also evaluated limited values for boosting efficacy and we did not examine vaccines with a half-life greater than 18 months. Third, our modelling scenarios are composed of archetypal seasonal transmission profiles and health system characteristics that broadly indicate the range of results for a particular prevalence setting. Moreover, we did not account for the heterogeneities in transmission or care-seeking likely to occur in endemic malaria settings. Previous comparisons between model estimates for geographic specific locations and setting archetypes do, however, show that estimates are similar for childhood vaccination with limited indirect benefits.(44) Lastly, vaccination coverage does not account for the nuances of access and uptake and only represents a broad metric of real-life implementation.

## Conclusions

The development of a highly efficacious, durable vaccine remains a priority for the malaria vaccine research and development community. Thus, obtaining an early understanding of a vaccine’s duration of protection and its efficacy decay profile is crucial. It is critical to incorporate an understanding of the duration of protection with appropriate clinical trial endpoints for burden reduction or infection prevention, alongside correlates of immunity. This will allow developers and stakeholders to assess and prioritize use cases aiming for greater public health benefits. Our modelling suggests that PEVs with high initial efficacy of more than 90% and a half-life duration of protective efficacy greater than 12 months offer opportunities for protection over multiple years, suggesting that yearly booster doses may not be needed, particularly in lower transmission settings. Our modelling results also provide a better understanding of trade-offs between vaccine performance properties, health system, and programmatic factors, and could support decision making for both clinical investment in and recommendations for new or next-generation malaria vaccines.

## Author contributions

JM and MAP conceived the study. JM conducted the analysis and wrote the first draft of the manuscript, MAP provided overall guidance and led development of the initial modelling framework. JM, LBM and NN were involved in updating the modelling framework for this study. All authors provided input, critically reviewed the manuscript and approved the final draft for submission.

## Data and code sharing

The datasets, code, and plot scripts used and analyzed in our study are all open access and available via the following link: https://doi.org/10.5281/zenodo.14012674.

## Data Availability

https://zenodo.org/records/14012674

## Acknowledgements

We would like to thank our colleagues in the Global Disease Modelling (GDM) group at The Kids Research Institute Australia, WA and the Disease Modelling research unit at the Swiss Tropical and Public Health Institute, University of Basel, Basel, for the support and discussions. We acknowledge and appreciate all the participants and investigators of the seasonal vaccination with RTS,S and seasonal chemoprevention trial in Mali and Burkina Faso. We would also like to acknowledge all participants of the 2021 ‘Malaria Prevention: Shaping Next-Gen Medical Interventions’ convening for their discussions and feedback. We also thank Jean-Luc Bodmer (Bill & Melinda Gates Foundation), Scott Miller (Gates Medical Research Institute), and Mary Hamel and Lindsay Wu (World Health Organization) for their support and discussions. Calculations were performed at the sciCORE Center for Scientific Computing at the University of Basel (http://scicore.unibas.ch/).

## Supplemental information

### 1. Materials and Methods

#### a. Mathematical transmission model

OpenMalaria is an individual based stochastic simulation model of malaria in humans linked to a deterministic model of malaria in mosquitoes.(1–3) The details on model structure and fitting process have been described in full.(4) The open-source code is available on (https://github.com/SwissTPH/openmalaria/wiki), including detailed documentation. At its core, OpenMalaria captures the clinical epidemiology and natural history of malaria and includes sub-models of infection in humans, blood stage parasite dynamics, infectiousness to mosquitoes and incidence of mortality and morbidity, and immunity. The model also accounts for genetic diversity, enabling the study of drug resistance, vaccine insensitivity, and vector susceptibility. All models have been fitted to field data from malaria endemic areas, as described in previous studies as shown in Table A. We provide an overview of OpenMalaria as used in our study.

In summary, malaria infection in humans is driven by the entomological inoculation rate (EIR) which affects the force of infection. Infection events and outcomes, parasite densities, mosquito and human elements are updated at each 5-day timesteps. Blood-stage parasite density depends on the time since infection and is affected by naturally acquired immunity. Acquired immunity, which reduces parasite density of subsequent infections, develops progressively following repeated exposure to infections and total parasitemia. Acute clinical illness depends on human host parasite densities and their pyrogenic threshold which evolves over time depending on previous exposure. Morbidity episodes can be uncomplicated or evolve to severe disease where a proportion of these severe cases leads to death. Transmission to mosquitoes depends on the parasite density present in the humans and a time-lag to allow the development of gametocytes. The overall infectiousness to mosquitoes is age-specific and has been validated to field data. OpenMalaria is also composed of a comprehensive simulation of the mosquito lifecycle and behavior towards human and animal hosts, including biting rates, resting and survival probabilities, embedded in a dynamic entomological model of the mosquito feeding cycle. All through the human and mosquito stages, the effect of various interventions on preventing infection, clinical episodes or onward transmission such as vector control, case management and vaccines can be modelled. The full characteristics of the model, modelled setting characteristics and entomological factors, outcome definitions are summarised in Table A and Table B.

**Table A:** Summary of OpenMalaria components and properties adapted from (5)

| Name | Description | References |
| --- | --- | --- |
| **Key epidemiological processes** |  |  |
| Malaria infection in humans | - Driven by the entomological inoculation rate (EIR) which is a model input and affects the force of infection in the simulated settings<br>- Human exposure to mosquitoes depends on age and is correlated with the body surface area | (6) |
| Progression of infections in humans: parasite densities and immunity | - Several sub-models of asexual parasite densities are included within OpenMalaria. In each model, blood-stage parasite density depends on the time since infection and is affected by naturally acquired immunity.<br>- Acquired immunity (both pre-erythrocytic and blood-stage) develops progressively following repeated exposure to infections and total parasitemia. Acquired immunity reduces parasite density of subsequent infections. | (1,6–8) |
|  | - The duration of infection follows a log-normal distribution and is estimated from a malaria therapy dataset.<br>- Super-infection (multiple distinct infections) is possible with cumulative parasite densities |  |
| Transmission from infected humans to mosquitoes | - Transmission to mosquitoes depends on the parasite density present in the humans (including a time-lag for gametocyte development)<br>- The age-specific contribution to overall infectiousness to mosquitoes was validated in against field data in malaria endemic countries. | (1,9,10) |
| Clinical illness, morbidity, and mortality | - Acute clinical illness depends on human host parasite densities and their pyrogenic threshold which evolves over time depending on the individual exposure history<br>- Acute morbidity episodes can be uncomplicated or evolve to severe episodes<br>- A proportion of the severe episodes leads to deaths | (1,11–13) |
| **Modelled characteristics of the transmission settings** |  |  |
| Population age structure | - Informed by health and demographic surveillance data from Ifakara, Tanzania | (11,14) |
| Transmission seasonality | - Seasonally forced, the same transmission pattern is reproduced each year in absence of interventions.<br>- Seasonal patterns are inputs to the model and users can define any patterns as needed e.g., perennial, one or two peak seasonal patterns, etc. | (6,15) |
| Entomological setting | - Comprehensive simulation of the mosquito lifecycle and behavior towards human and animal hosts (biting, resting) embedded in a dynamic entomological model of the mosquito feeding cycle<br>- Multiple vector species can be simulated simultaneously | (3) |
| Case Management | - Modelled through a comprehensive decision tree-based model which determines the corresponding treatment implications depending on the occurring clinical events such as fevers and seeking of care<br>- Its representation includes specification of access to official or nonofficial care, access to hospital for severe cases, diagnostic tests (use, specificity, sensitivity, and threshold of detection), treatments for first, second line and non-official care, effects of treatment, case fatality rate, case sequelae and cure rates | (16) |
| **Modelled interventions** |  |  |
| Vector control | - Vector interventions can deter mosquitoes from humans, reduce the probability that a mosquito bites a protected human host or reduce the probability that a mosquito survives host feeding.<br>- These include bed nets, indoor residual spraying, repellents e.t.c | (17–19) |
| Drugs and Vaccines | - Drugs and vaccines act at different stages of the parasite life cycle (transmission blocking, anti-infective, blood-stage clearance) and their action is defined by their initial efficacy, half-life, and decay | (20,21) |
| Deployment characteristics | - Interventions can be deployed for several rounds to a targeted group of individuals and specified coverages (proportion of the population reached with the intervention)<br>- Interventions can be deployed by enrolling individuals into cohorts and tracking cohort outcomes, facilitating clinical trial simulation |  |
| **Simulation regimens and model variants** |  |  |
| Time Steps | - Simulation outputs are tracked every 5 days (one time step) |  |
| Model variants | - Varying assumption on treatment, immunity decay, heterogeneity of transmission etc results in 14 model variants. In the current study, we only use the base model. | (22) |
| **Software availability and documentation** |  |  |
| Source code and wiki page available on GitHub: https://github.com/SwissTPH/openmalaria/ |  |  |

**Table B:** Simulated scenarios and outcome measures.

| Parameter | Description |
| --- | --- |
| **Primary Outcomes** |  |
| Infection prevalence reduction | The relative reduction in infection prevalence compared with a no intervention counterfactual, where prevalence was defined as the proportion of all malaria infections (detectable by a highly sensitive diagnostic such as PCR) across the six-, 12-, or 24-month period after the final vaccine booster dose in the fifth year, or in six-monthly intervals until 18 months after primary series doses |
| Clinical incidence reduction | The relative reduction in the incidence of clinical cases compared with a no intervention counterfactual, where incidence was defined as the number of new, uncomplicated malaria cases across the six-, 12-, or 24-month period after the final vaccine booster dose in the fifth year, or in six-monthly intervals until 18 months after primary series doses<br>Uncomplicated malaria was defined as an episode of symptomatic malaria, detectable by rapid diagnostic test with 94.2% specificity and a detection limit of 50 parasites per microlitre, where symptoms did not qualify as severe malaria. |
| Severe malaria cases reduction | The relative reduction in the incidence of severe malaria cases compared with a no intervention counterfactual, where severe cases of malaria were evaluated across the six-, 12-, or 24-month period after the final vaccine booster dose in the fifth year, or in six-monthly intervals until 18 months after primary series doses.<br>An episode of severe malaria was an episode of hospitalized symptomatic malaria, with symptoms qualifying as severe malaria or with co-morbidities |
| **Target population** | Vaccine impact was evaluated for two target populations<br>- Children under five years old<br>- Children under three years old for multi-seasonal impact in the first and second years after vaccination |
| **Setting characteristics** |  |
| *PfPR*<sub>2-10</sub> | Baseline parasite prevalence before intervention of between 10% and 50% |
| Seasonality | Highly seasonal (4-month profile), Seasonal (6-month profile) and constant transmission |
| Mosquito species | *Anopheles gambiae* |
| **Health system characteristics** |  |
| Access to care | Probability of accessing effective curative malaria treatment within 14-days of symptomatic malaria. Treatment is assumed to clear all blood stage infections within 5 days<br>- Low probability of 10%<br>- Moderate probability of 30%<br>- High probability of 45% |
| Diagnostic | Rapid diagnostic tests (RDTs) |

#### b. Vaccine deployment in settings with constant transmission

**Fig A:**
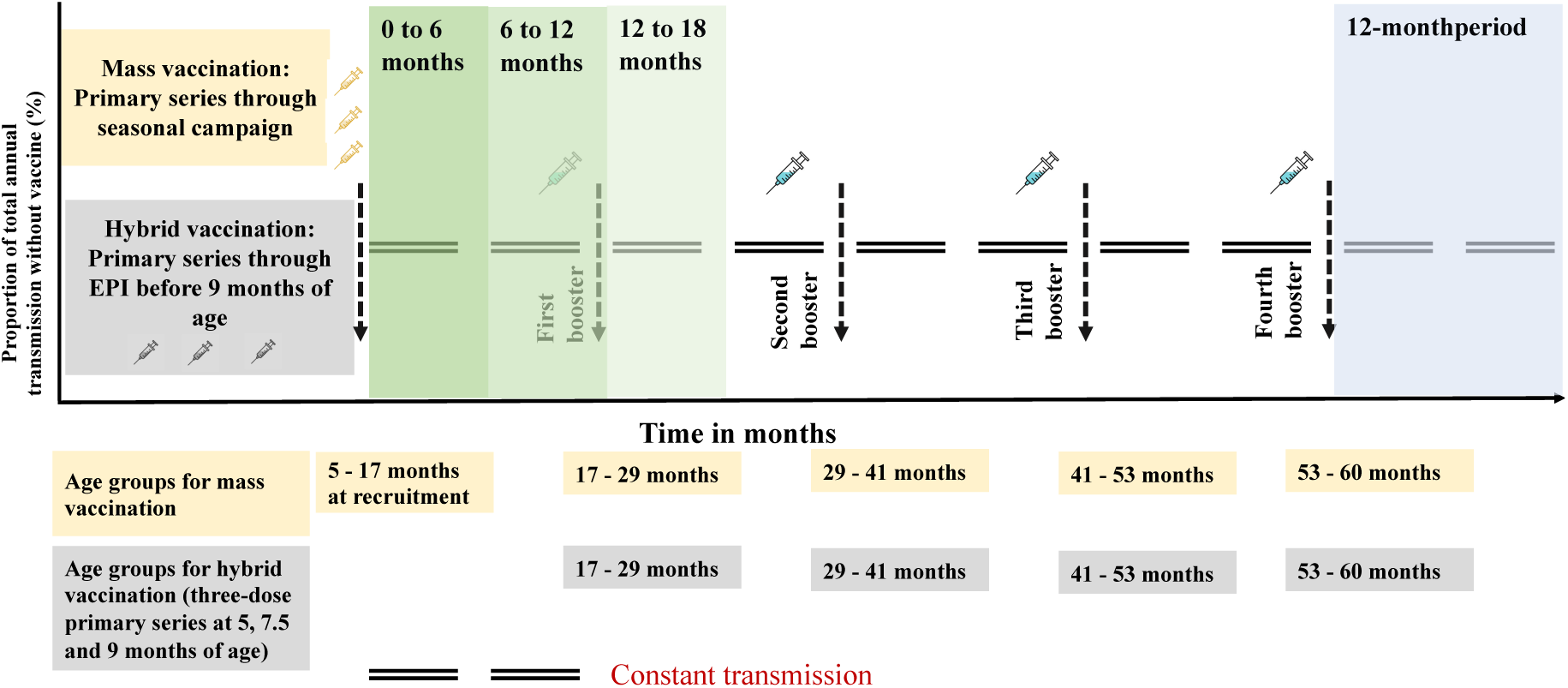
Schematic illustration of the simulated vaccine deployment schedules for a five-year vaccination program shown for a setting with constant transmission. Illustration shows the five-year vaccination program with a primary series dosage, the timing of the annual boosters and the age groups at each dose for both vaccine delivery schedules, hybrid vaccination (grey boxes) and mass vaccination (brown boxes). For the mass vaccination schedule, the primary series and annual booster doses are deployed before the peak transmission season. For the hybrid vaccination schedule, doses for the primary series are deployed continuously as part of an age-based immunization schedule while the annual boosters are also deployed before the peak transmission season. The green shading shows the zero to 18 months period where the multi-seasonal or multi-year vaccine impact is evaluated by comparing a cohort of children who received the primary series doses and the first booster (dose four) and those who only received the primary series doses. The blue shading shows the 12-month period following the final annual booster dose (dose seven) in the fifth year where the public health vaccine impact is evaluated for children who received the primary series followed by annual boosters.

### 2. Validation of vaccine and chemoprevention properties using clinical trial data

#### a. Trial characteristics

We used results from a recently conducted Phase 3 clinical trial for seasonal vaccination with RTS,S in Mali and Burkina Faso to validate the vaccine properties of RTS,S. Specifically, we validated the vaccine’s efficacy reached after boosting in the two years following primary vaccination. This comparison informed model assumptions around the efficacy profile of a first booster/fourth dose given 12 months after the three-dose primary series, rather than 18 months after, as implemented in the original Phase 3 clinical trial conducted in seven sites in African countries and a second booster/fifth dose given 24 months after. We validate the results for the clinical trial arms, including: seasonal vaccination using RTS,S alone, seasonal vaccination using RTS,S given in combination with seasonal chemoprevention using SPAQ, and seasonal chemoprevention using SPAQ alone. Similarly, using a similar approach described and the results from the same Phase 3 clinical trial in Mali and Burkina Faso we calibrate the preventive half-life duration of seasonal chemoprevention using SPAQ.

We compared our model results across two seasonal transmission profiles with seasonality patterns taken from Mali and Burkina Faso (Fig B). We selected the baseline EIR translating to a *PfPR_2-10_* between 5-65% and *PfPR_6-12_* between 10-70% to reproduce the malaria epidemiology in the two study sites. We also modelled other interventions including vector control. Each trial participant was given a long-lasting insecticide at the beginning of the trial. Any other interventions not reported as part of the trial were assumed to remain constant and are reflected in the baseline EIR. We assume a 45% probability of accessing curative malaria treatment within 14 days of the onset of symptoms. We reproduce the clinical trial by recruiting a cohort of children aged five to 17 months and following them up for two years, as done in the clinical trial.

In a second analysis evaluating the duration of protection in the aforementioned seasonal chemoprevention and vaccination trial in Mali and Burkina Faso, the 6-monthly vaccine protective efficacy amongst children receiving SMC was at 75.9% [95% CI 67.0–82.4%] in the first year after the three-dose primary vaccination series, 63.6% [95% CI 57.2–69.1%] in the second year after the first booster/fourth dose, and 60.1% [95% CI 52.9–66.2%] in the third year after the second booster/fourth dose. This analysis showed wider confidence intervals after the initial six to seven months (∼200 days) following vaccination in each year reflecting a biphasic decay profile with a long tail.(23,24) This translates to between 10% to 30% drop in protective efficacy against clinical disease in years two and three compared with year one, in both countries.(23,24)

**Table C:**
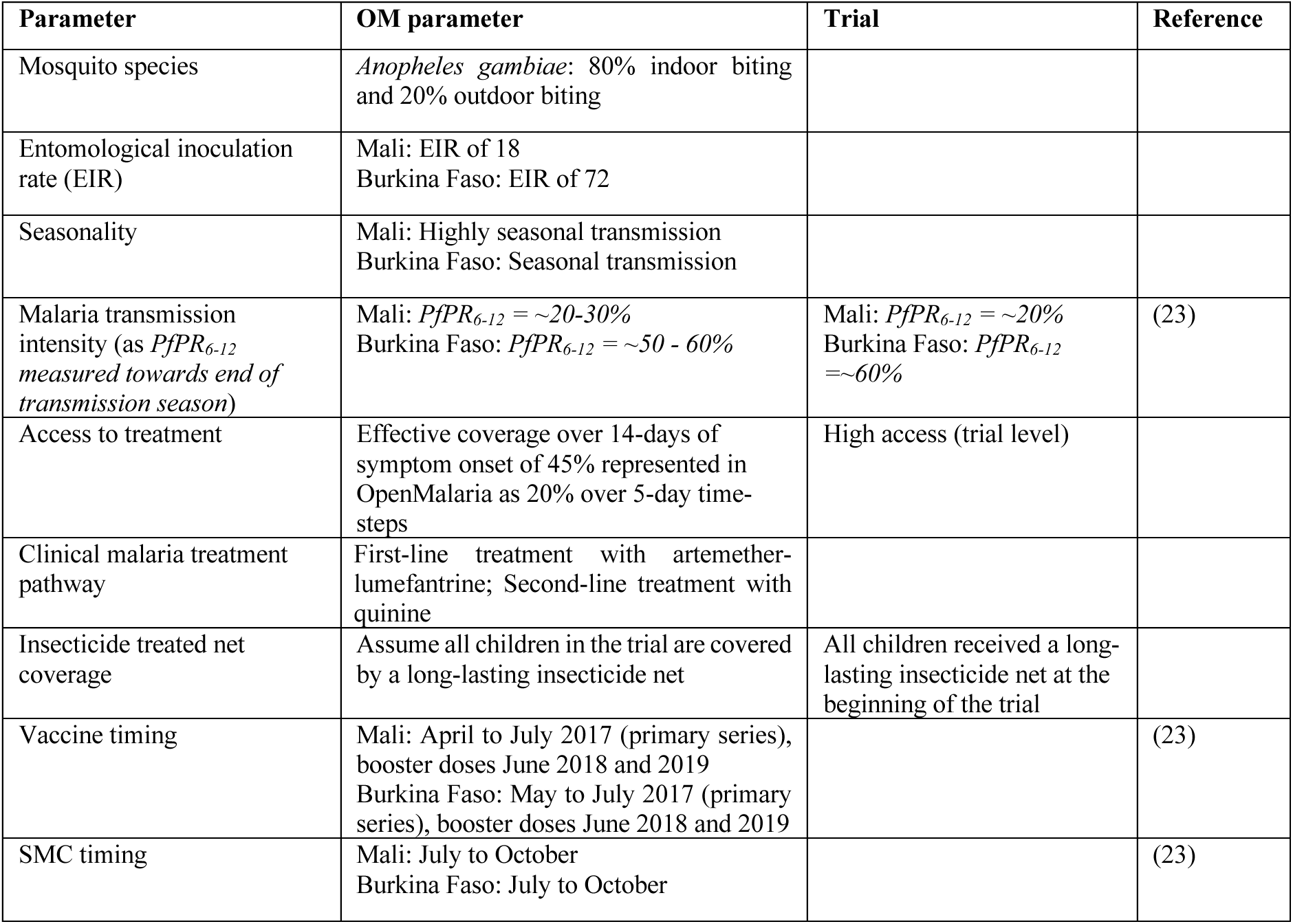
Trial characteristics matched to modelled input to check intervention parameterization.

#### b. Vaccine decay profile and seasonality profiles

**Fig B:**
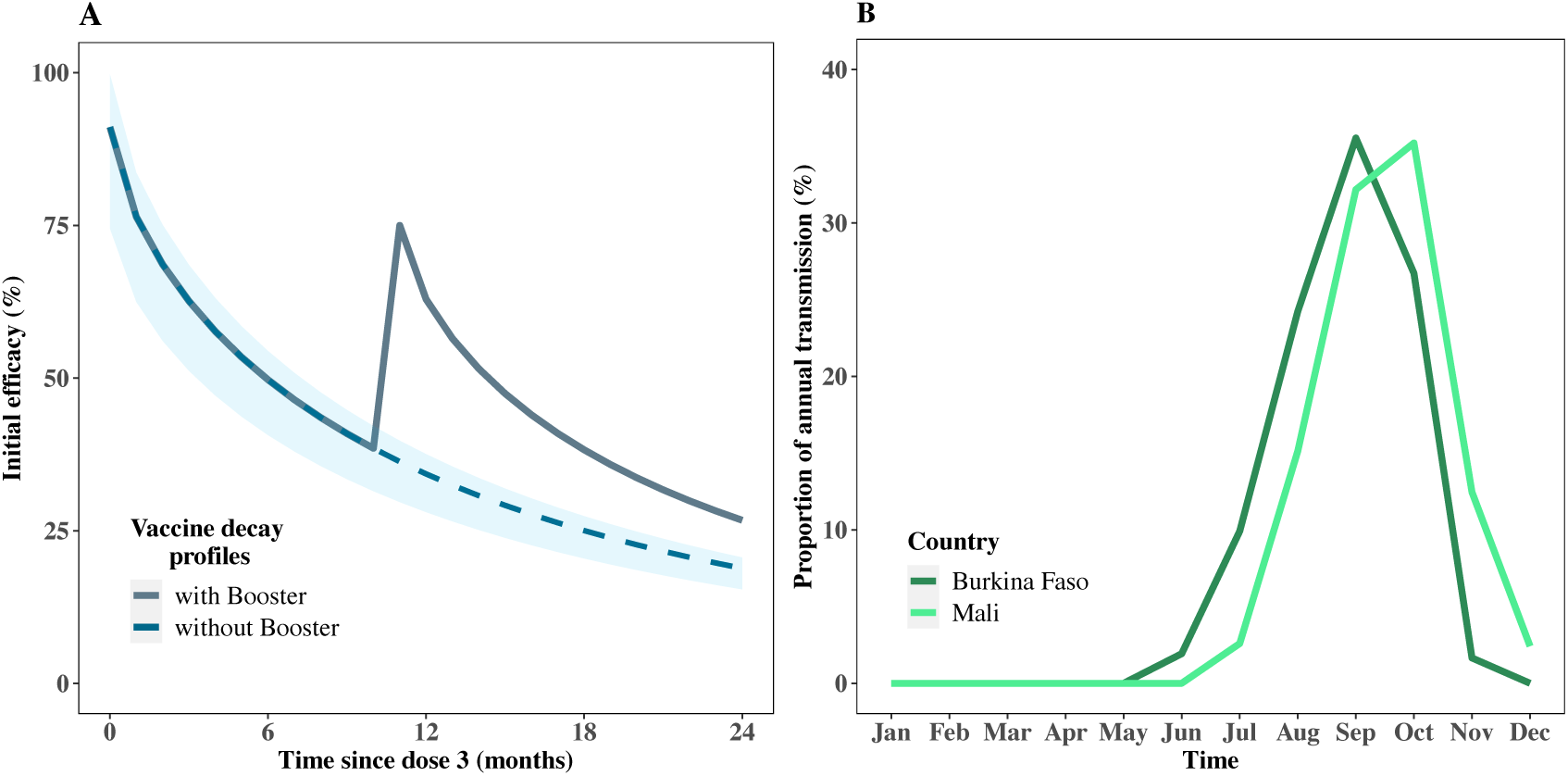
Schematic illustration of vaccine decay profile over two years and the yearly seasonal profiles used as model input for Burkina Faso and Mali. A) Illustration shows a two-year vaccine efficacy decay profile following the three dose primary series and the first annual booster given in the first year. The initial efficacy reached after dose three is 91.1%, a half-life duration of 7.32 months with a biphasic decay with shape parameter k = 0.69. The booster dose is assumed to follow the same decay profile. The light grey-blue solid line represents the booster, a dashed darker grey-blue line represents the scenario without booster. B) Yearly seasonal profiles used as model inputs for Burkina Faso and Mali.

#### c. Modelled scenarios and trial endpoints

Modelled estimates from the original Phase 3 clinical trial conducted in seven sites in African countries,(26,27) show RTS,S to have a high initial vaccine efficacy of 91.1% [95% CI 74.5– 99.7%] following the three-dose primary vaccination, with a biphasic immunity decay profile where the shape parameter was k = 0.69, and a half-life duration (time until initial efficacy decays by 50%) was 7.32 months [95% CI 6–10 months] for children aged 5-17 months at recruitment. Vaccine efficacy could be restored to 49.9% [95% CI 32.0–68.6%] following a single booster dose given 18 months after the three-dose primary vaccination.(26)

In this study, the previously reported estimates for initial efficacy after primary vaccination, decay function and half-life duration of protection are used as model inputs. To estimate the vaccine initial efficacy reached after the first and second boosters, we assume that the boosting efficacy falls by between 10% to 20% after the first booster/fourth dose, and 20% to 30% after the second booster/fifth dose. Assumptions on parameter ranges, modelled for boosting efficacy and the decline in protection during the second and third years in this study, were based on previous clinical and modelling evidence for RTS,S, (23–25,27) as described above. Vaccine efficacy decays with a biphasic exponential function,(25,28) declining rapidly in the initial six to seven months after the primary vaccination series followed by a slow decay with a long tail. The same decay profile is assumed for the primary series and following the booster doses.

To match the clinical trial endpoints, we first extracted the monthly clinical incidence over the three-year trial period and we used in the model fitting. We also calculated the cumulative incidence and the hazard ratios for both countries separately and combined to reproduce the published estimates.(23) For this modelling study, we only focus on the primary outcome as reported from the clinical trial: uncomplicated clinial episodes.

#### d. Bayesian optimization approach

We used a bayesian optimization (BO) approach, to fit our model results to clinical trial data, providing the best fit values for both countries for all the three trial arms. BO is an approach used for hyperparameter optimization in machine learning to find the global optima with a limited number of function evaluations.(30) BO combines a probabilistic model of the objective function, in this case a Gaussian Process (GP) model, with an acquisition function to minimize a loss function. For this study, we integrated BO with PyTorch and the botorch library(30), a framework for deep learning which allowed for efficient computation.(29) We used the residual sum of squares (RSS) as the loss function to ensure improved regression model accuracy. The RSS is a measure of the discrepancy between observed and the predicted values, in this case the clinical trial data and modelled results, and is defined as:

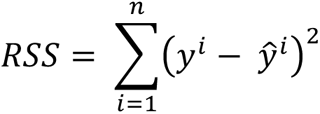

We used a GP as the surrogate model to provide a probabilistic estimate of the objective function and used the Expected Improvement (EI) as the acquisition function.(29,31) EI quantifies the expected improvement over the current best observation guiding the search towards regions with a high probability of improvement, hence balancing exploration of uncertainty and exploitation of the most promising points to evaluate next. The function returns the best candidate observations given the mean and variance of the surrogate model. The implementation began with initializing a set of training points with synthetic data used to fit the initial GP model. During each iteration, the EI acquisition function was optimized to identify the most likely points for evaluation in the subsequent step. These points were then evaluated using the RSS and the outcomes added to the training set, hence, updating the GP model iteratively. For each trial arm, the process was replicated for 1000 iterations progressively refining the search space for the optimal parameters that minimize the RSS.

#### e. Bayesian optimization results for all arms in the two countries

**Table D:** Optimization results for all parameters in the two countries across the three trial arms.

| Country | Mali |  |  | Burkina Faso |  |  |
| --- | --- | --- | --- | --- | --- | --- |
| Arm | Vaccine alone arm | SMC alone arm | Combined arm | Vaccine alone arm | SMC alone arm | Combined arm |
| EIR | 18 | 18 | 18 | 72 | 72 | 72 |
| Access | 45% | 45% | 45% | 45% | 45% | 45% |
| Initial Efficacy dose 4 | 0.87998 |  | 0.85391 | 0.80507 |  | 0.80507 |
| Initial Efficacy dose 5 | 0.65950 |  | 0.76480 | 0.80000 |  | 0.80000 |
| SMC (SPAQ half-life duration of protection) |  | 20.38441 | 26.92995 |  | 20.81110 | 20 |
| SMC (SPAQ decay of protection, Weibull parameter (k)) |  | 5.28359 | 4.28407 |  | 2.54896 | 4.30773 |

| Other interventions | ITN | ITN | ITN | ITN | ITN | ITN |
| --- | --- | --- | --- | --- | --- | --- |

#### f. Additional in silico modelling results

**Fig C:**
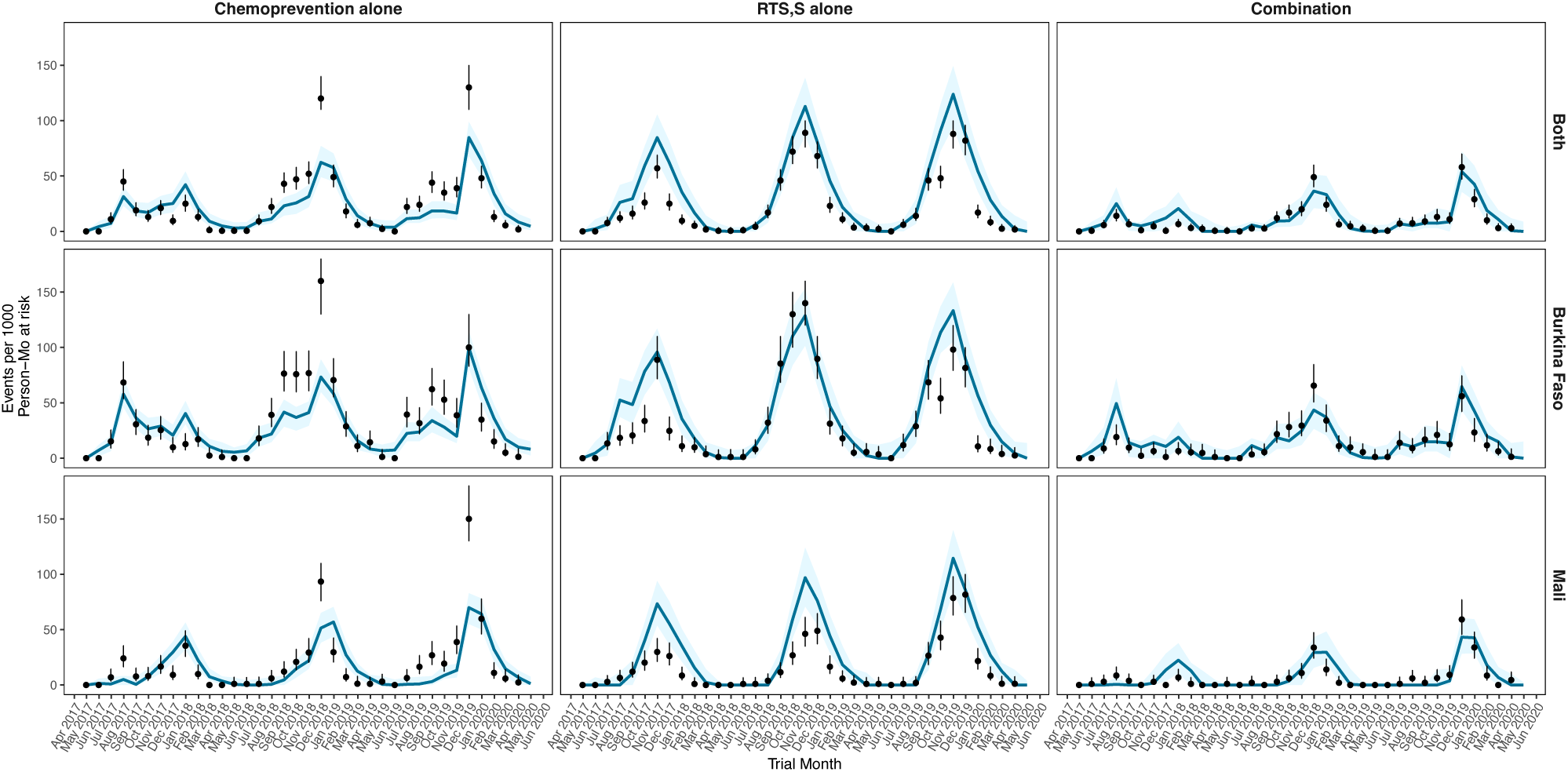
Monthly clinical incidence for the clinical trial data (black dots) compared with model simulations (blue lines/triangles) using the best fit assumption for the efficacy of annual booster doses four and five and SPAQ preventive half-life duration. Best fit for Burkina Faso (*Pf*PR_6-12_ is 50−60% and seasonal transmission), Mali (*Pf*PR_6-12_ is 20−30% and highly seasonal transmission) and both countries combined shown for the three trial arms. The black dots shown with 95% confidence intervals represent the trial field data and the blue lines/triangles illustrate the modelled output from the simulations with the shaded region showing the confidence intervals averaged over 100 seeds. In this figure, the parameters were optimized for the arm where the vaccine and chemoprevention were combined and used to predict the model results for the arms where chemoprevention or vaccination were given alone.

**Fig D:**
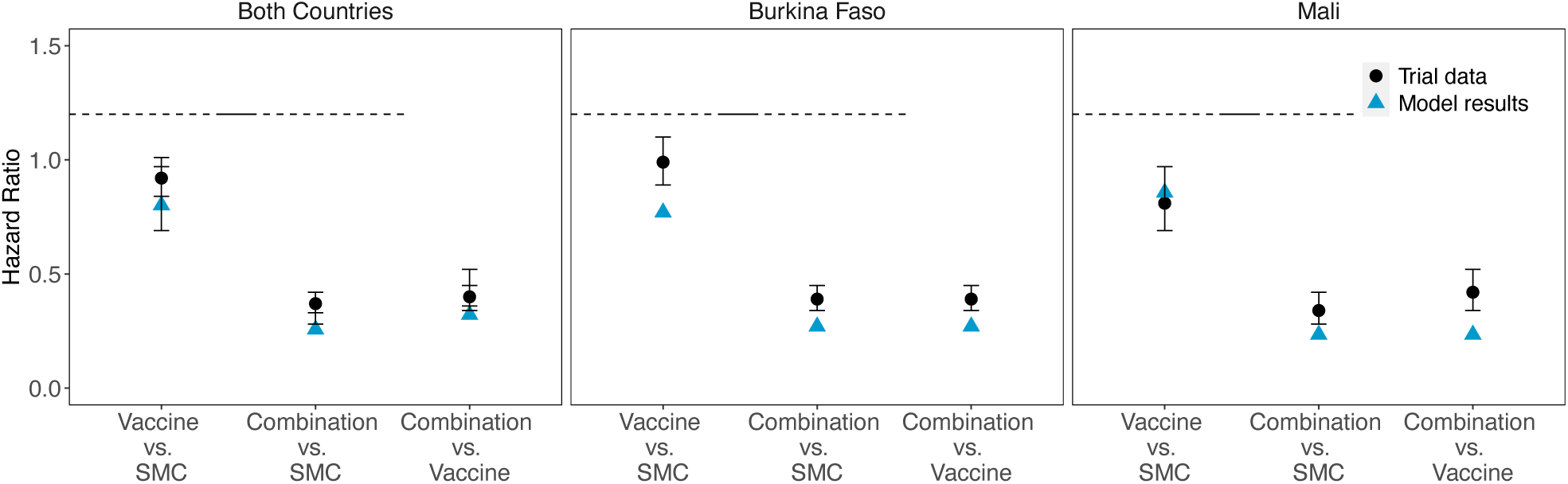
Hazard Ratios the clinical trial data (black dots) compared with model simulations (blue lines/triangles) using the best fit assumption for the efficacy of annual booster doses four and five and SPAQ preventive half-life duration. Hazard ratios for the three trial arms in Burkina Faso (*Pf*PR_6-12_ is 50−60% and seasonal transmission), Mali (*Pf*PR_6-12_ is 20−30% and highly seasonal transmission) and both countries combined. The black dots shown with 95% confidence intervals represent the trial field data and the blue lines/triangles illustrate the modelled output from the simulations with the shaded region showing the confidence intervals averaged over 100 seeds. In this figure, the parameters were optimized for the arm where the vaccine and chemoprevention were combined and used to predict the model results for the arms where chemoprevention or vaccination were given alone.

### 3. Additional modelling results

#### a. Public health impact of improved PEVs on the malaria burden

**Fig E:**
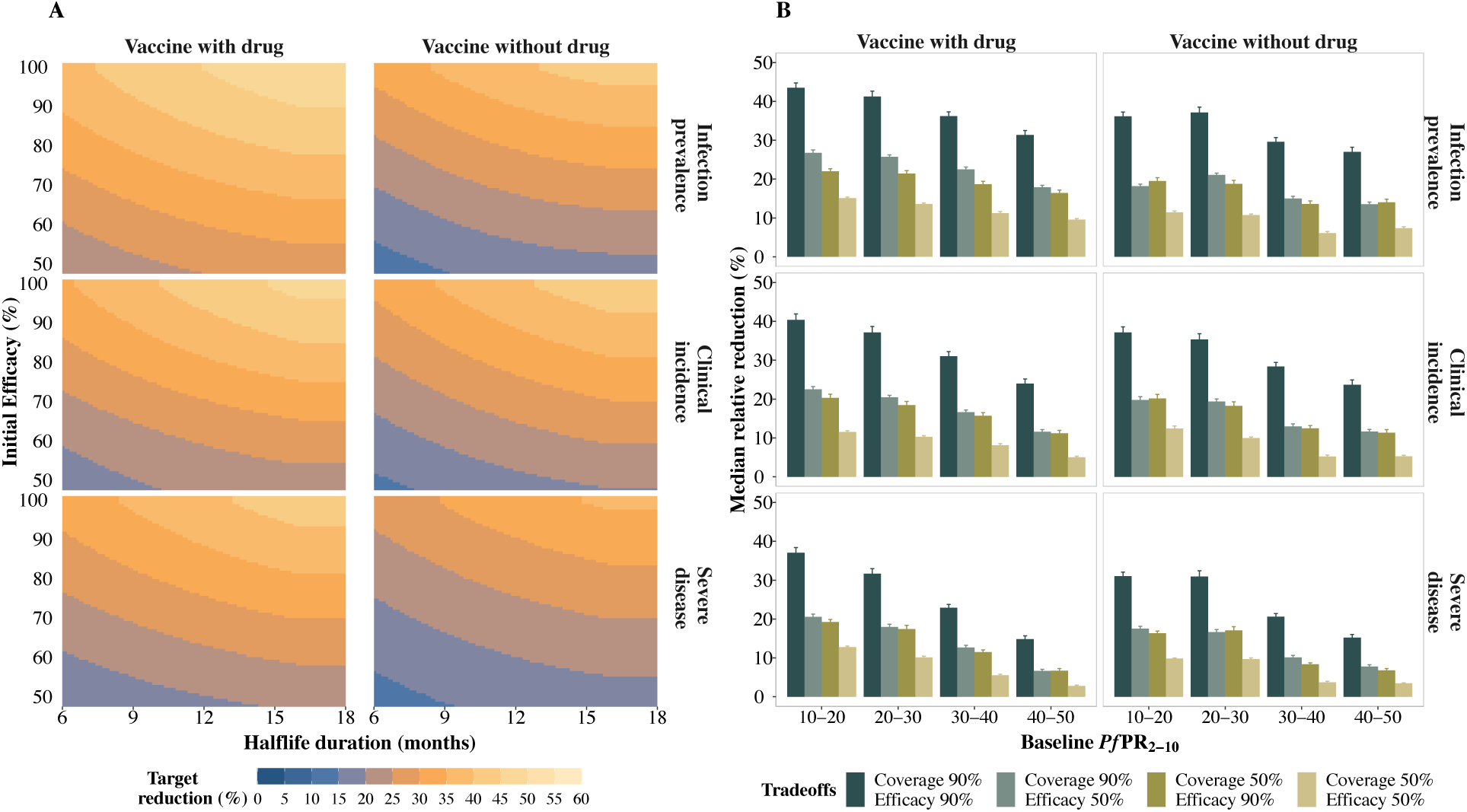
Predicted relative burden reduction in the 12-month period following the final annual booster dose, compared with a no-intervention counterfactual. A) Target reduction (%) in infection prevalence (top row), clinical incidence (middle row), and severe disease (bottom row) illustrating trade-offs between initial efficacy and half-life duration of protection in settings where baseline *Pf*PR_2-10_ ranged between 10% and 20%. The initial efficacy ranged from 50% to 100%, half-life duration from six to 18 months, and assuming a primary series vaccination coverage of 90%. B) Median (interquartile range (IQR)) relative reduction in infection prevalence (top row), clinical incidence (middle row), and severe disease (bottom row), considering varying levels of baseline *Pf*PR_2-10_, coverage, and initial efficacy for a long duration vaccine with a half-life duration between 12 and 18 months. Results are shown for a setting where the PEV was not co-administered with a blood stage clearance drug, for a four-month short seasonality profile, for the mass vaccination deployment schedule, in settings with a 30% probability of accessing curative treatment within 14 days of symptom onset.

**Fig F:**
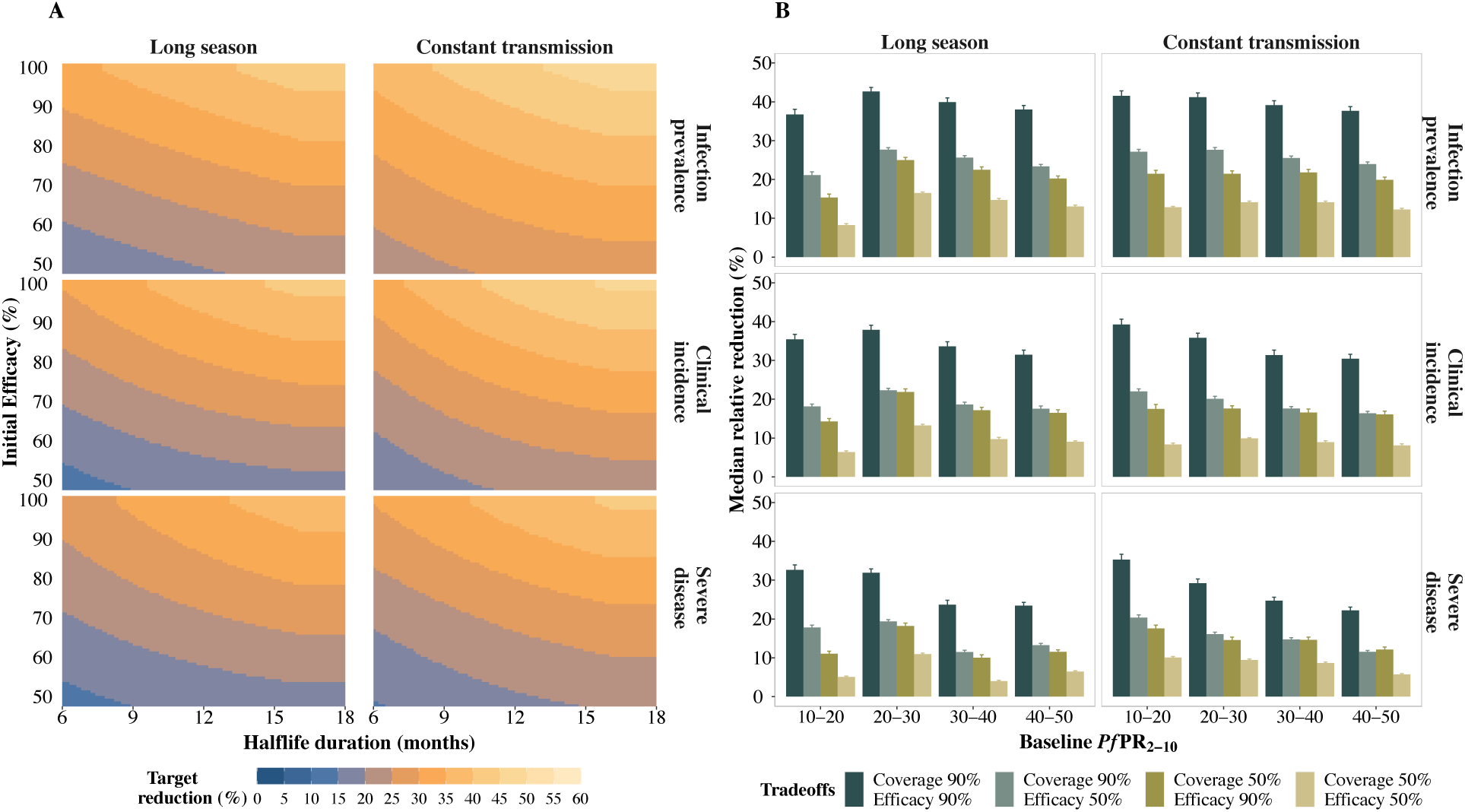
Predicted relative burden reduction in the 12-month period following the final annual booster dose, compared with a no-intervention counterfactual. A) Target reduction (%) in infection prevalence (top row), clinical incidence (middle row), and severe disease (bottom row) illustrating trade-offs between initial efficacy and half-life duration of protection in settings where baseline *Pf*PR_2-10_ ranged between 10% and 20%. The initial efficacy ranged from 50% to 100%, half-life duration from six to 18 months, and assuming a primary series vaccination coverage of 90%. B) Median (interquartile range (IQR)) relative reduction in infection prevalence (top row), clinical incidence (middle row), and severe disease (bottom row), considering varying levels of baseline *Pf*PR_2-10_, coverage, and initial efficacy for a long duration vaccine with a half-life duration between 12 and 18 months. Results are shown where the PEV was co-administered with a blood stage clearance drug, for a 6-month long seasonality profile and constant transmission, for the mass vaccination deployment schedule, in settings with a 30% probability of accessing curative treatment within 14 days of symptom onset.

**Fig G.**
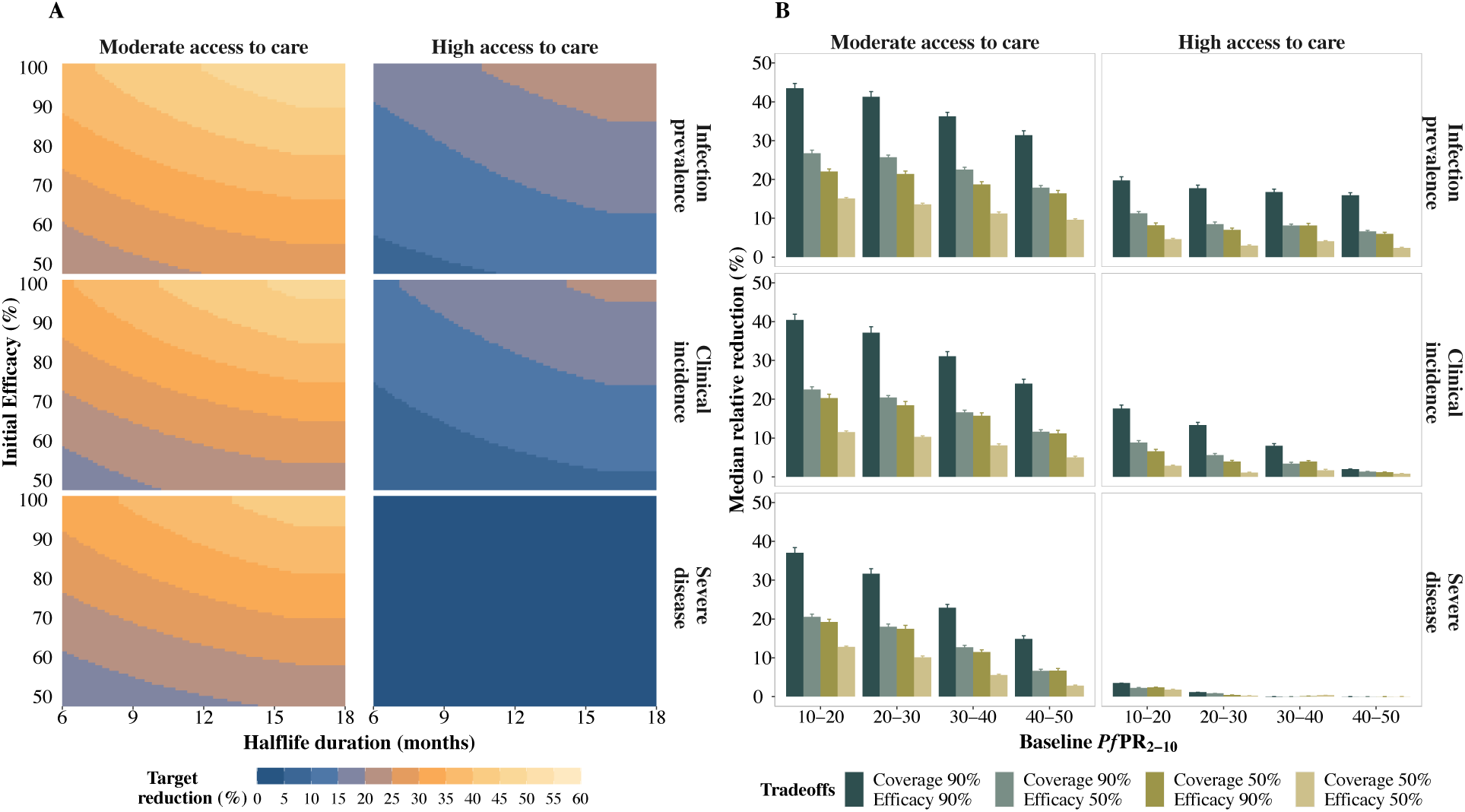
Predicted relative reduction in infection prevalence, clinical incidence and severe disease for the 12-month period following the final annual booster dose, compared with a no-intervention counterfactual. A) Target reduction (%) in infection prevalence (top row), clinical incidence (middle row), and severe disease (bottom row) illustrating trade-offs between initial efficacy and half-life duration of protection in settings where baseline *Pf*PR_2-10_ ranged between 10% and 20%. The initial efficacy ranged from 50% to 100%, half-life duration from six to 18 months, and assuming a primary series vaccination coverage of 90%. B) Median (interquartile range (IQR)) relative reduction in infection prevalence (top row), clinical incidence (middle row), and severe disease (bottom row), considering varying levels of baseline *Pf*PR_2-10_, coverage, and initial efficacy for a long duration vaccine with a half-life duration between 12 and 18 months. Results are shown for a PEV co-administered with a blood stage clearance drug, for a four-month short seasonality profile, for the mass vaccination deployment schedules, in settings with a moderate (30%) and high (70%) probability of accessing curative treatment within 14 days of symptom onset.

**Fig H:**
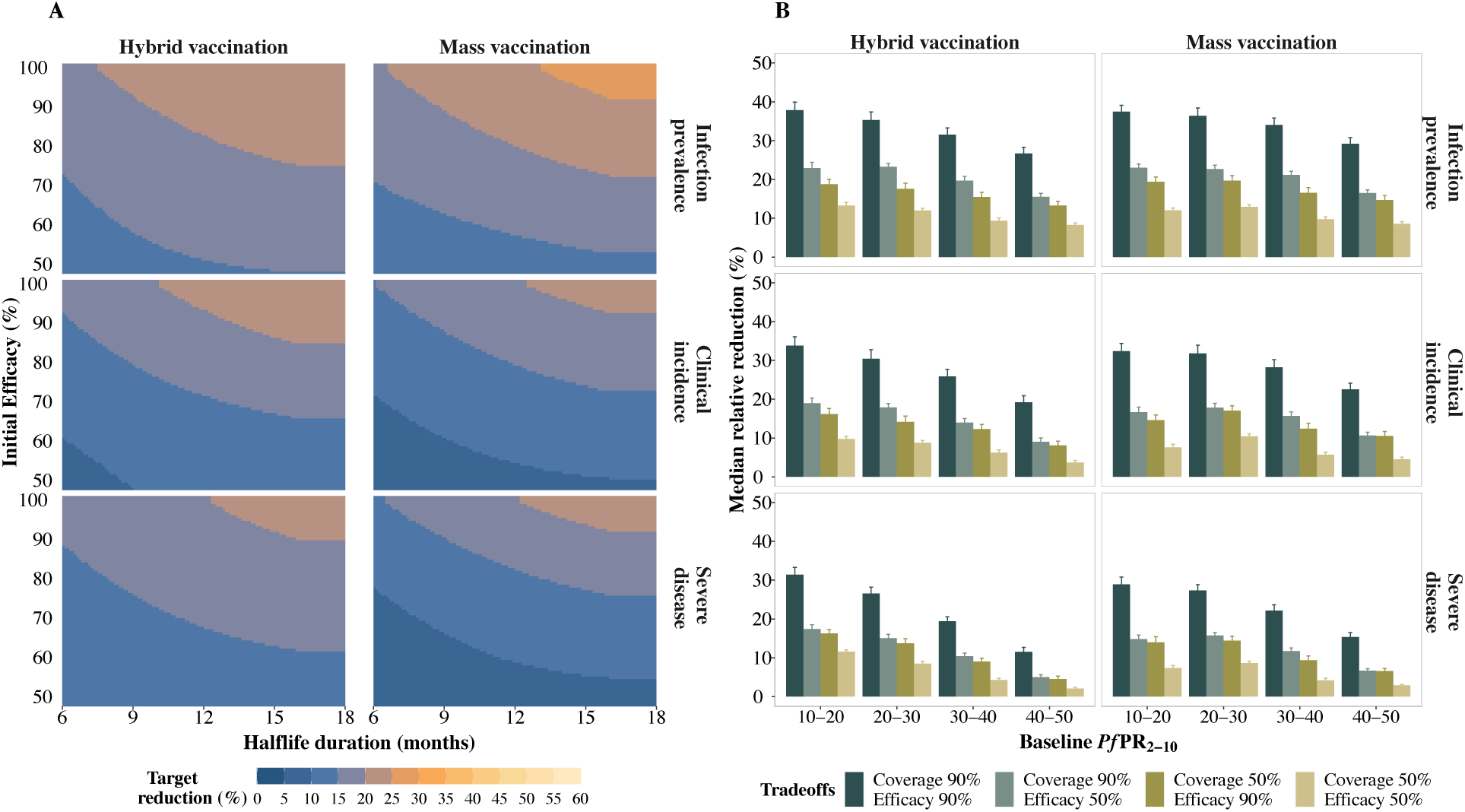
Predicted relative reduction in infection prevalence, clinical incidence and severe disease for the 12-month period following the final annual booster dose, compared with a no-intervention counterfactual. A) Target reduction (%) in infection prevalence (top row), clinical incidence (middle row), and severe disease (bottom row) illustrating trade-offs between initial efficacy and half-life duration of protection in settings where baseline *Pf*PR_2-10_ ranged between 10% and 20%. The initial efficacy ranged from 50% to 100%, half-life duration from six to 18 months, and assuming a primary series vaccination coverage of 50%. B) Median (interquartile range (IQR)) relative reduction in infection prevalence (top row), clinical incidence (middle row), and severe disease (bottom row), considering varying levels of baseline *Pf*PR_2-10_, coverage, and initial efficacy for a short duration vaccine with a half-life duration between six and 12 months. Results are shown for a PEV co-administered with a blood stage clearance drug, for a four-month short seasonality profile, for both deployment schedules, in settings with a 30% probability of accessing curative treatment within 14 days of symptom onset.

#### b. Drivers of impact following a five-year vaccination program

**Fig I:**
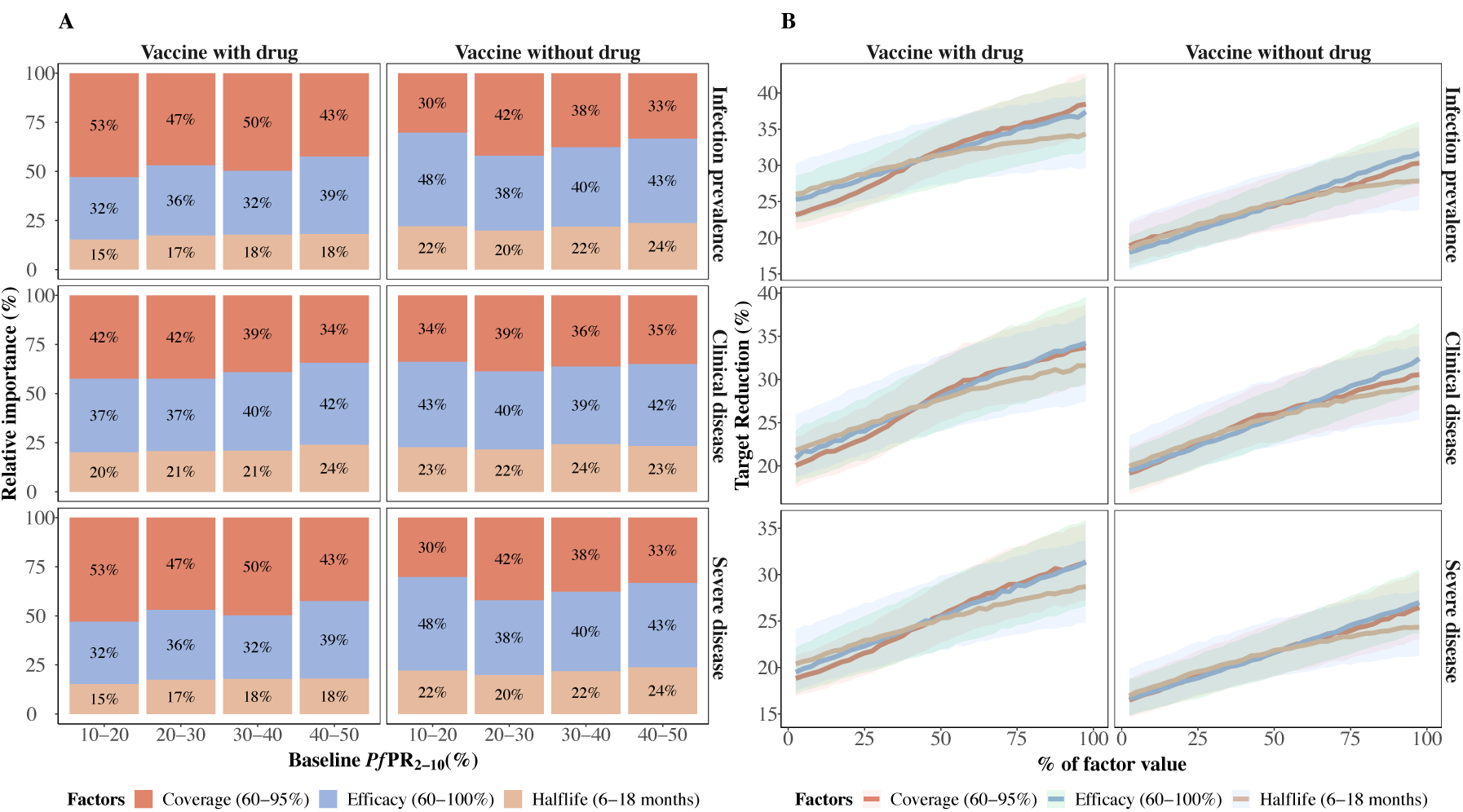
Factors influencing vaccine impact on predicted burden reductions for the 12-month period following the final annual booster dose compared with a no-intervention counterfactual. A) Bars indicate the total Sobol effect indices, which explain the variance in predictions of relative reduction. These indices can be interpreted as the proportion of variation in the outcome attributed to changes in each variable. Results are shown across various baseline *Pf*PR_2-10_ values and span different parameter ranges for initial efficacy (70% to 100%), half-life duration (six to18 months), and vaccination coverage (60% to 90%). B) The influence of the impact-driving factors on predicted reductions in infection prevalence and clinical incidence for settings where *Pf*PR_2-10_ lies between 20% and 30%. The different lines and shaded areas depict the median and interquartile range (IQR) of proportional contribution, as estimated through global sensitivity analysis over the variable parameter ranges for initial efficacy (60% to 100%), half-life duration (six to 18 months) and vaccination coverage of (60% to 95%). Results are shown for a setting where the PEV was not co-administered with a blood stage clearance drug, for a four-month short seasonality profile, for the mass vaccination deployment schedules, in settings with a 30% probability of accessing curative treatment within 14 days of symptom onset.

**Fig J:**
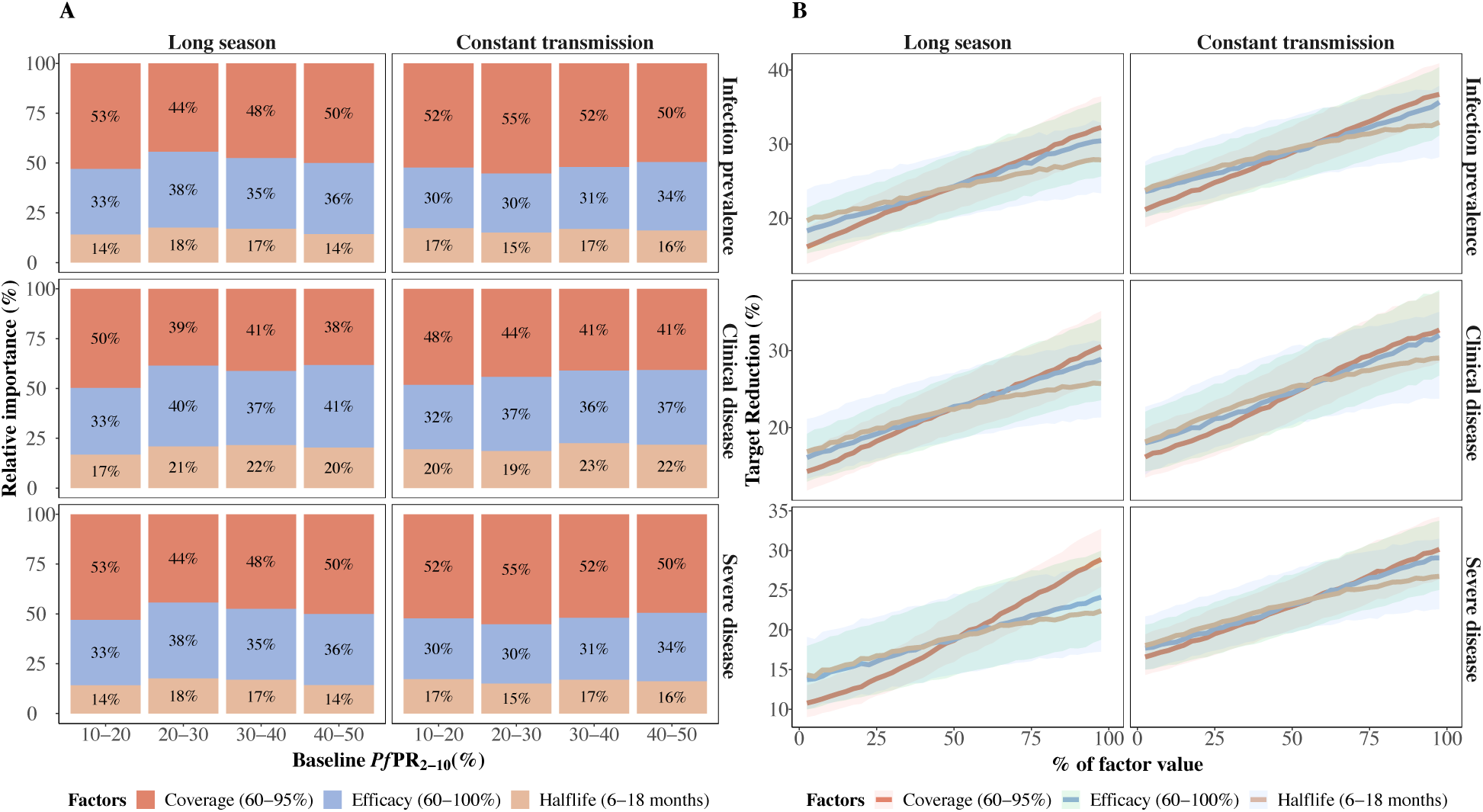
Factors influencing vaccine impact on predicted burden reductions for the 12-month period following the final annual booster dose compared with a no-intervention counterfactual. A) Bars indicate the total Sobol effect indices, which explain the variance in predictions of relative reduction. These indices can be interpreted as the proportion of variation in the outcome attributed to changes in each variable. Results are shown across various baseline *Pf*PR_2-10_ values and span different parameter ranges for initial efficacy (70% to 100%), half-life duration (six to18 months), and vaccination coverage (60% to 90%). B) The influence of the impact-driving factors on predicted reductions in infection prevalence and clinical incidence for settings where *Pf*PR_2-10_ lies between 20% and 30%. The different lines and shaded areas depict the median and interquartile range (IQR) of proportional contribution, as estimated through global sensitivity analysis over the variable parameter ranges for initial efficacy (60% to 100%), half-life duration (six to 18 months) and vaccination coverage of (60% to 95%). Results are shown where the PEV was co-administered with a blood stage clearance drug, for a 6-month long seasonality profile and constant transmission, for the mass vaccination deployment schedule, in settings with a 30% probability of accessing curative treatment within 14 days of symptom onset.

**Fig K:**
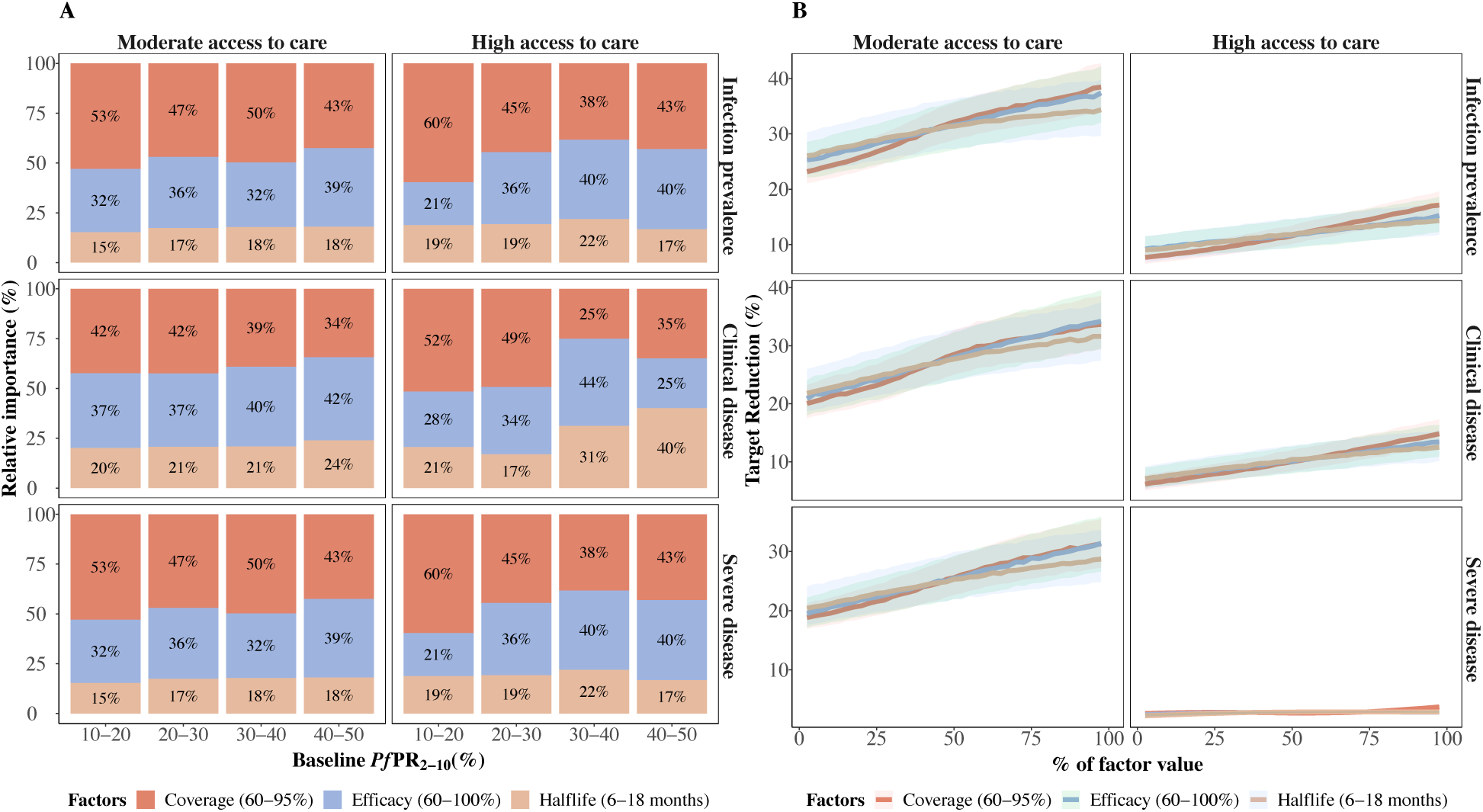
Factors influencing vaccine impact on predicted burden reductions for the 12-month period following the final annual booster dose compared with a no-intervention counterfactual. A) Bars indicate the total Sobol effect indices, which explain the variance in predictions of relative reduction. These indices can be interpreted as the proportion of variation in the outcome attributed to changes in each variable. Results are shown across various baseline *Pf*PR_2-10_ values and span different parameter ranges for initial efficacy (70% to 100%), half-life duration (six to18 months), and vaccination coverage (60% to 90%). B) The influence of the impact-driving factors on predicted reductions in infection prevalence and clinical incidence for settings where *Pf*PR_2-10_ lies between 20% and 30%. The different lines and shaded areas depict the median and interquartile range (IQR) of proportional contribution, as estimated through global sensitivity analysis over the variable parameter ranges for initial efficacy (70% to 100%), half-life duration (six to 18 months) and vaccination coverage of (60% to 90%). Results are shown for a PEV co-administered with a blood stage clearance drug, for a four-month short seasonality profile, for the mass vaccination deployment schedules, in settings with a moderate (30%) and high (70%) probability of accessing curative treatment within 14 days of symptom onset.

#### c. Multi-year vaccine impact in the two years following primary vaccination

**Fig L:**
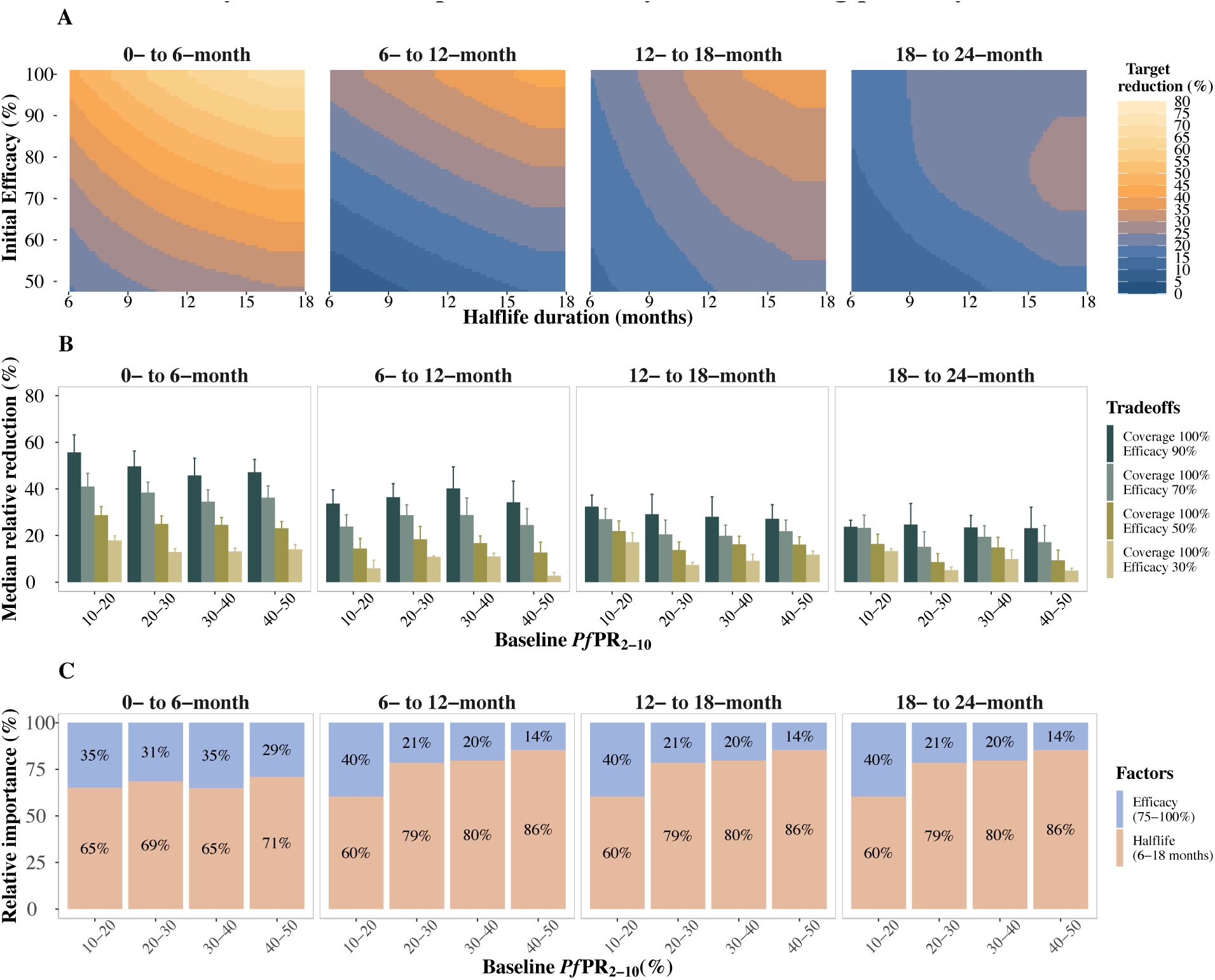
Predicted relative reduction and impact-drivers on clinical incidence in the zero to 18 months following primary vaccination for children who did not receive a booster compared to a no-intervention counterfactual. A) Median (interquartile range (IQR)) relative reduction in clinical incidence for different levels of *Pf*PR_2-10_, two levels of initial efficacy (50%, 95%), for a long duration vaccine with a half-life between 12 and 18 months and primary series vaccination coverage of 100%. B) Trade-offs between initial efficacy and half-life duration of protection for clinical incidence reduction in settings where baseline *Pf*PR_2-10_ ranged between 10% and 20%, initial efficacy ranged from 50% to 100%, and half-life duration from six to 18 months, assuming a primary series vaccination coverage of 100%. C) Bars indicate the total Sobol effect indices, which explain the variance in predictions of relative reduction in clinical incidence. These indices can be interpreted as the proportion of variation in the outcome attributed to changes in each variable. Results are shown over different parameter ranges for initial efficacy (75% to 100%) and half-life duration (six to18 months), where vaccination coverage was fixed at 100%. Results are shown for a PEV co-administered with a blood stage clearance drug, for the zero- to six-, six- to 12- and 12- to 18-month periods following primary vaccination, for a six-month short seasonality profile, the hybrid vaccination schedule, in settings with a 30% likelihood of accessing curative treatment within 14 days of symptom onset.

**Fig M:**
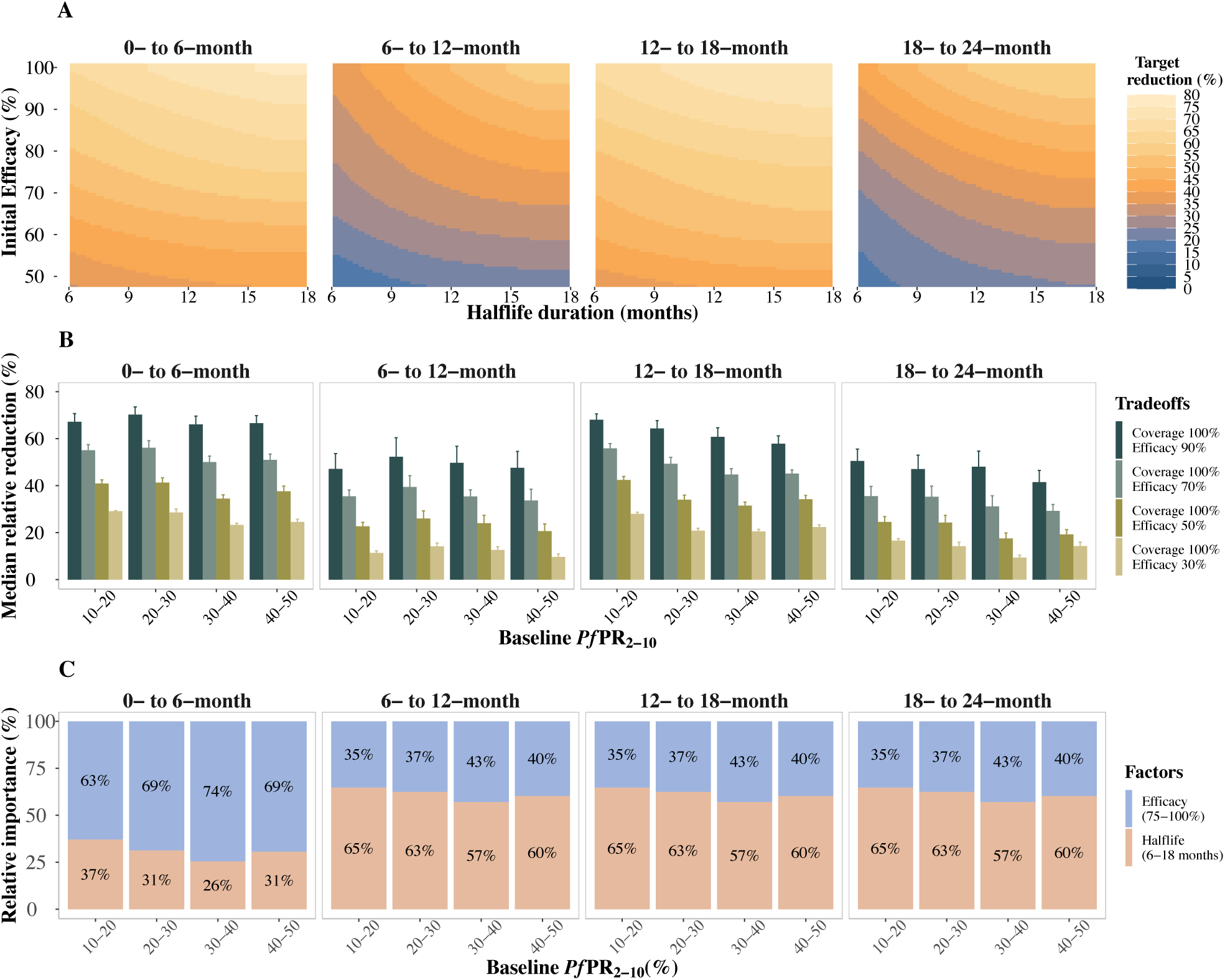
Predicted relative reduction and impact-drivers on clinical incidence in the zero to 18 months following primary vaccination for children who received a booster compared to a no-intervention counterfactual. A) Median (interquartile range (IQR)) relative reduction in clinical incidence for different levels of *Pf*PR_2-10_, two levels of initial efficacy (50%, 95%), for a long duration vaccine with a half-life between 12 and 18 months and primary series vaccination coverage of 100%. B) Trade-offs between initial efficacy and half-life duration of protection for clinical incidence reduction in settings where baseline *Pf*PR_2-10_ ranged between 10% and 20%, initial efficacy ranged from 50% to 100%, and half-life duration from six to 18 months, assuming a primary series vaccination coverage of 100%. C) Bars indicate the total Sobol effect indices, which explain the variance in predictions of relative reduction in clinical incidence. These indices can be interpreted as the proportion of variation in the outcome attributed to changes in each variable. Results are shown over different parameter ranges for initial efficacy (75% to 100%) and half-life duration (six to18 months), where vaccination coverage was fixed at 100%. Results are shown for a PEV co-administered with a blood stage clearance drug, for the zero- to six-, six- to 12-, 12- to 18- and 18- to 24-month periods following primary vaccination, for a six-month short seasonality profile, for the mass vaccination schedule, in settings with a 30% likelihood of accessing curative treatment within 14 days of symptom onset.

**Fig N:**
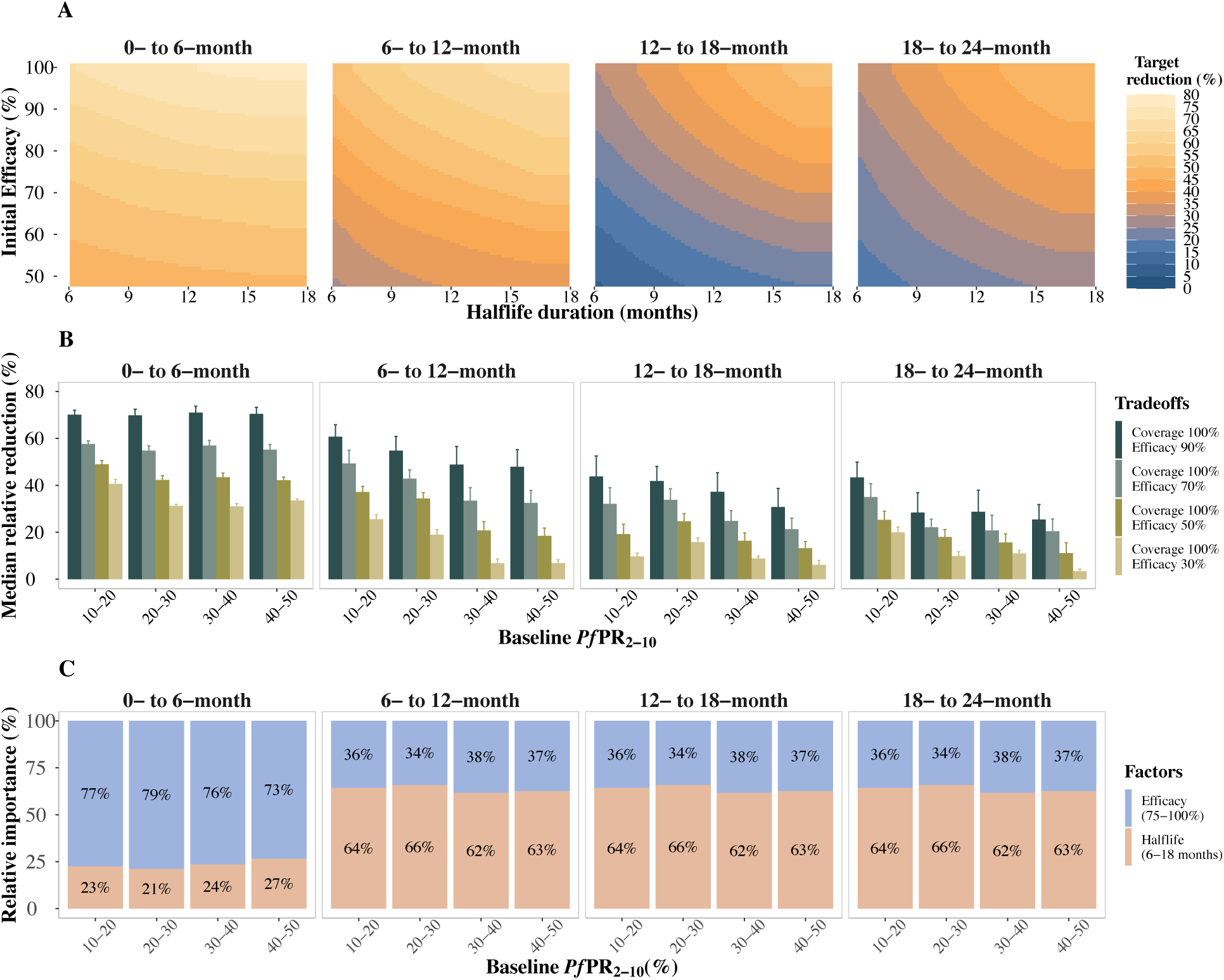
Predicted relative reduction and impact-drivers on clinical incidence in the zero to 18 months following primary vaccination for children who did not receive a booster compared to a no-intervention counterfactual. A) Median (interquartile range (IQR)) relative reduction in clinical incidence for different levels of *Pf*PR_2-10_, two levels of initial efficacy (50%, 95%), for a long duration vaccine with a half-life between 12 and 18 months and primary series vaccination coverage of 100%. B) Trade-offs between initial efficacy and half-life duration of protection for clinical incidence reduction in settings where baseline *Pf*PR_2-10_ ranged between 10% and 20%, initial efficacy ranged from 50% to 100%, and half-life duration from six to 18 months, assuming a primary series vaccination coverage of 100%. C) Bars indicate the total Sobol effect indices, which explain the variance in predictions of relative reduction in clinical incidence. These indices can be interpreted as the proportion of variation in the outcome attributed to changes in each variable. Results are shown over different parameter ranges for initial efficacy (75% to 100%) and half-life duration (six to18 months), where vaccination coverage was fixed at 100%. Results are shown for a PEV co-administered with a blood stage clearance drug, for the zero- to six-, six- to 12-, 12- to 18- and 18- to 24-month periods following primary vaccination in the mass vaccination deployment schedule, for a setting with constant transmission, in settings with a 30% likelihood of accessing curative treatment within 14 days of symptom onset.

